# Quantifying heat exposure and its related mortality in Rio de Janeiro City: evidence to support Rio’s recent heat protocol

**DOI:** 10.1101/2025.01.17.25320740

**Authors:** João Henrique de Araujo Morais, Débora Medeiros de Oliveira e Cruz, Valeria Saraceni, Caroline Dias Ferreira, Gislani Mateus Oliveira Aguilar, Oswaldo Gonçalves Cruz

**Affiliations:** Sergio Arouca National School of Public Health, Oswaldo Cruz Foundation (Fiocruz), Rio de Janeiro - RJ, Brazil; Rio de Janeiro Municipal Health Secretariat (SMS), Rio de Janeiro - RJ, Brazil; Medical Education Institute (IDOMED), Rio de Janeiro - RJ, Brazil; Program of Scientific Computing (PROCC), Oswaldo Cruz Foundation (Fiocruz), Rio de Janeiro - RJ, Brazil

**Keywords:** extreme heat, mortality, epidemiology, elderly, brazil

## Abstract

Under a climate change scenario, extreme heat episodes show an increase in frequency and intensity, with a scaling impact in Latin American cities. Recently, Rio de Janeiro City developed its heat protocol, using the amount of hours spent over specific heat thresholds as its trigger metric. This study gathers mortality data by 17 different causes in Rio, in a 12.5 year period (2012-2024). We use Distributed Lag Non-Linear Models (DLNM) to assess the relationship between different heat exposure metrics to mortality among the young (< 65) and elderly (>=65y), including a novel metric called Heat Area Above a Threshold (HAAT). In the study period, there were 466,121 deaths in the city from natural causes. Deaths due to diabetes, hypertensive diseases, Alzheimer’s/dementia, renal failure and even undetermined deaths were strongly associated with extreme heat episodes, especially among the elderly. The proposed HAAT metric showed better performance on explaining mortality for most causes (10 out of the 17), when compared to temperature or heat index, or commonly used heat wave definitions. The results dialogue with Rio’s heat protocol, evaluating the cut-off points defined and proposing simpler definitions using the HAAT metric. An exposure to a HAAT of 64°C*h increases mortality by natural causes by 50%, and 91.2°C*h already doubles the mortality risk. Main strengths of the study lie on the comparison of different heat exposure metrics and the investigation of cause-specific mortality in a period when recent and remarkable heat waves occurred. There is still fragility when considering a compound index such as the Heat Index, and social and spatial differences on heat-related mortality should also be considered in future models. The proposed metric, however, appears as a relevant indicator to distinguish unusually warm days that lead to elevated mortality, and could guide definitions for Heat Warning Systems.

## Introduction

As the planet reaches its warmest years in 2023 and 2024^1,2^, heat wave episodes are becoming more frequent and intense across the globe ^3–6^. This temperature increase and a considerable portion of its related mortality has been shown to be due to human action ^7,8^ and projections show that extreme events such as heat waves will get longer and even more frequent in the future ^8,9^. The health burden caused by heat is well documented in the literature. Exposure to extreme temperatures can affect the pathophysiological mechanisms of thermoregulation, distribution of body flow and sweating mechanisms ^10,11^. Studies around the world have extensively shown associations of heat waves with cardiovascular and respiratory diseases ^12–15^, renal diseases ^16–18^, mental health ^4,19^ and pregnancy outcomes ^20,21^. Certain population groups are known to be at increased risk, like older populations and those with coexisting health conditions ^11,22–25^. Disparities in risk are also shown to exist across gender and race ^4,26^.

There has been an increasing attention to heat effects in health in Latin American countries ^15,27^, as the Latin population is aging ^28^ and the number of heat-related extreme episodes in the region is scaling ^8,29^. In Brazil, most single-city studies concentrate in the cities of São Paulo ^23,30,31^ and Rio de Janeiro ^24,32^. Recently, Moraes *et al*. ^23^ showed a higher risk for cardiovascular and respiratory mortality under different definitions of heat waves among the elderly population in São Paulo. Silveira and collaborators showed similar results for the city of Rio de Janeiro ^24^ and also for 32 municipalities in the Brazilian Amazon ^33^. Santos *et al.* ^4^ showed an increase in the number, intensity and duration of heatwaves in the 14 most populous metropolitan regions of Brazil, and estimated 48,075 heat-related excess deaths in the period of 2000-2018.

As cities have dealt with heat waves for some time, heat warning systems (HWS) have been implemented around the world over the last decades ^34–37^, with the goal of warning, communication and protection of the population. Brazil faced its worst heat wave in history in November 2023 ^38^, which resulted in a remarkable fan’s death due to heat exhaustion during a concert in Rio de Janeiro ^39^. In June 2024, Rio launched a protocol for dealing with extreme heat in the city ^40^.

The existing HWS from other countries employ a range of metrics for trigger warning ^36,37^, from bio-meteorological indices, such as Heat Index ^41^ or Humidex ^42^ to thermal stress indices like Universal Thermal Climate Index ^43^ or Excess Heat Factor ^44^. Rio’s protocol, that establishes five heat risk levels (NC1 - “normality” to NC5 - “extreme heat danger”), differs from others since it is based on the amount of hours spent in a day above a certain Heat Index (HI) threshold, in consecutive warm days. The cut-off points defined for the heat levels: 4 hours with HI >= 36°C (levels NC2 and NC3), 4 hours with HI >= 40°C (level NC4) and 2 hours with HI >= 44°C (NC5) were not based on heat-mortality associations. The present study intends, then, to explore heat-mortality associations for 17 different groups of causes of death in Rio de Janeiro city (RJC), in two population age-groups, varying the metric of heat exposure: average daily temperature and heat index, and the amount of hours of heat exposure. Finally, it proposes an index that summarizes the period of exposure to high heat in a given period - the Heat Area Above a Threshold (HAAT), and compares how it can help explain mortality in unusually warm days and potentially guide heat protocols based on the amount of heat exposure.

## Methods

### Data and variables

#### Mortality data

Mortality data was retrieved from the Brazilian Mortality Information System (*Sistema de Informações de Mortalidade* - SIM), referring to deaths that occurred from 2012 to the first six months of 2024. We filtered only deaths of residents of RJC (CO_MUN_RES = 330455). The ICD-10 field of underlying cause of death was used to classify deaths into 17 groups, based on 13 groups specified in a CDC’s Excess Mortality Study ^45^, along with additional groups for urinary tract deaths, undetermined deaths, selected deaths (all of the previously included) and natural causes of death (all deaths except external causes). The groups are summarized in Table 1. Daily death counts by cause group were aggregated, and divided into two age-groups: young (0-64 years old) and elderly (65+).

**Table 1.** Causes of deaths groups included in the study, and respective ICD-10 codes. Based on ^45^.

| Cause of death group | ICD-10 codes |
| --- | --- |
| Influenza and Pneumonia | J09-J18 |
| Chronic lower respiratory diseases | J40-J47 |
| Other diseases of the respiratory system | J00–J06, J20–J39, J60–J70, J80–J86, J90–J96, J97–J99, R09.2, U04 |
| Hypertensive diseases | I10-I15 |
| Ischemic heart disease | I20-I25 |
| Heart failure | I50 |
| Cerebrovascular diseases | I60-I69 |
| Other disease of the circulatory system | I00–I09, I26–I49, I51, I52, I70–I99 |
| Malignant neoplasms | C00-C97 |
| Alzheimer disease and dementia | G30, G31, F01, F03 |
| Diabetes | E10-E14 |
| Renal Failure | N17-N19 |
| Sepsis | A40-A41 |
| Urinary Tract Disorder | N39 |
| Undetermined causes | R00-R99 |
| Selected causes | All of the above |
| Natural causes | All except external causes (S00-Y99) |

#### Climate data

Temperature (T) and relative humidity (H) data were retrieved for 16 weather stations, from 3 different sources: Alerta Rio System (7 stations), Brazilian National Meteorological Institute (*Instituto Nacional de Meteorologia* - INMET, 4 stations) and Aeronautics Command Meteorology Network (*Rede de Meteorologia do Comando da Aeronáutica* - REDEMET, 5 stations). Stations locations are represented in Supplementary Figure 1. Heat Index as described in Steadman ^41^ was calculated for all stations (when data for T and H were available), and the hourly median heat index was considered for the analysis. This is the procedure already done in Rio’s heat monitoring, in an attempt to obtain a summary metric for the municipality, disregarding extreme values from specific stations ^40^.

For each day in the study period (Jan/2012 - Jun/2024), the following variables were calculated:

- Average daily temperature (T_med_), calculated as a mean of the median hourly temperature among the stations;
- Average daily heat index (HI_med_), using the same principles as T_med_;
- Amount of hours in the day on which it was registered a median Heat Index above a certain threshold. Threshold values varied from HI = 32°C to HI = 44°C;
- The Heat Area Above a Threshold (HAAT), a proposed metric to represent the amount of exposure to high values of heat index in a given day.

#### The HAAT metric

The HAAT is calculated as the area between the hourly observed heat index and the horizontal line *HI*_*med*_ = *HI*_*thresh*_:

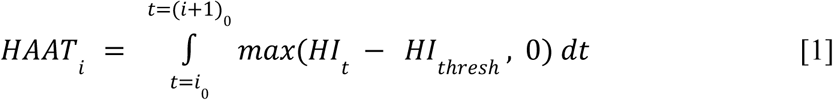

*HAAT_i_* is the HAAT value for a given day i, i_0_ is the first hour of day i, (i+1)_0_ is the first hour of day i+1. HI_t_ is the heat index value for moment t, and HI_thresh_ is the defined base threshold. In this study, we considered two thresholds: HI_thresh_=36°C, to match the first threshold defined in RJC heat protocol; and HI_thresh_= 32°C, to compare whether a broader limit could explain mortality any better. We also considered multiple day accumulated HAAT, with the hypothesis that continuous exposure to heat in consecutive days could lead to worse mortality excess. Therefore, the HAAT values included in the study were:

- Daily HAAT, and accumulated HAAT for 3, 5 and 7 days for HI_thresh_ = 32°C;
- Daily HAAT, and accumulated HAAT for 3, 5 and 7 days for HI_thresh_ = 36°C;

Common heat wave (HW) definitions in the literature were also calculated for the study period, in order to compare whether HAAT could explain mortality better than existing HW definitions. The definitions included were: Daily T_med_ higher than historical quantiles Q90, Q92.5, Q95 and Q97.5 for 2, 3 and 4 days.

### Statistical Analyses

Daily mortality outcomes were modeled using Generalized Additive Models (GAMs), with crossbasis functions from Distributed Lag Non-Linear Models (dlnm) ^46^. Exploratory analyses were done to identify whether day of week (dow) was an important confounder to consider in the models. Other relevant variables for environment-related mortality, like air quality (PM_10_ and PM_2.5_) were not included due to low coverage available for the city of Rio.

#### Model structure

The base model structure accounts for mortality long-term trends and yearly seasonality. Since mortality patterns changed relevantly during the Covid-19 pandemic ^47^, a specific term was included to account for this change during that period.

For 16 of the death cause groups, a Poisson distribution was used since no overdispersion was detected. The natural causes group was the only group identified with overdispersion, for which a negative binomial distribution was chosen. Therefore,

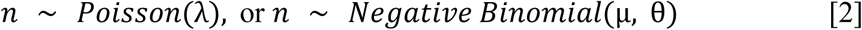

where n is the daily death count for a given cause and age group. The expected value is then modeled as:

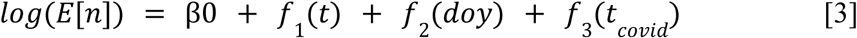

where:

- *f*_1_ is the smoothing function to capture the overall trend of death counts over the years. A thin plate spline was used with k=12 knots (one for each complete year in the analysis);
- *f*_2_ captures the yearly seasonality of deaths, based on the day of the year (*doy*, *doy* ɛ [1, 366]). For f_2_ a cubic cyclic spline was used, with k=6 knots.
- - f_3_ is the function used to capture the differential pattern of death counts during the period with most Covid-19 burden. For Rio de Janeiro, this period was defined from March 1st, 2020, first month after the arrival of Covid-19 in Brazil; to December 17th, 2022, date when the last significant Covid-19 hospitalizations wave ended ^48^. Instead of using a dummy variable to represent the differences in this period, a continuous index was used, in order to capture the heterogeneity of the Covid-19 effect on the death pattern inside this time window. Therefore, the index t_covid_, upon which f_3_ is applied, is represented as:

*t*_*covid*_ ɛ [1, …, 1022], in the period when Covid-19 effect was relevant: t_covid_ = 1 for March 1st, 2020; and t_covid_ = 1022 for December 17th, 2022 (time difference of 1022 days).
*t*_*covid*_ = 0 otherwise (outside of this time period).
For this term a cubic cyclic spline was also used, k=8 knots.

To better understand each term in the model structure, Supplementary Figure S1 illustrates what each term captured in a model for selected causes of death, among all age groups. A similar model, but without the Covid-19 term and using a gaussian likelihood, was fitted to T_med_ and HI_med_, to visually compare its exceedance patterns with the ones from mortality data.

#### Distributed lag non linear models

The base model defined above was extended iteratively to include the exposure variables of interest, using the crossbasis function from R package dlnm to account for lagged effects ^49^.

##### Models for temperature and heat index

Models using T_med_ and HI_med_ were fitted to understand the general relation between the 17 outcome groups and heat exposure in RJC, and also to evaluate whether HI_med_ would perform as a better predictor than T_med_ for mortality, since this relation is not certain in the literature ^50,51^. For these two exposure variables, a B-spline was considered in the crossbasis function, using 3 degrees of freedom (df). The spline choices for each variable were defined after a sensitivity analysis, detailed later on.

##### Models for amount of hours of exposure

The attempt to include hours of exposure to heat as an explanatory variable in the models had the goal of evaluating the thresholds of 4 hours of HI >= 36° C, 4 hours of HI >= 40° C and 2 hours of HI >= 44°C, used for changing heat levels in RJC’s heat protocol. Models using the amount of hours over each threshold (from 32°C to 44°C) as a continuous variable were fitted, using a natural spline with 1 df.

##### Models for HAAT

The HAAT variables were fitted similarly using penalized splines with df = 15. A combination of models using both HAAT and a continuous daily average metric (T_med_ or HI_med_) was also tested, in order to identify whether days with high HAAT have an added effect on mortality and if so, compare them with the added effects of heat waves, described subsequently.

##### Models for heat wave definitions and added effects

In order to compare the performance of the proposed HAAT variable with heat wave (HW) definitions used in the literature, models with varying HW definitions were included. For each definition, a model with HW only as an explanatory variable and with HW + T_med_ were fitted. HW was included in the crossbasis function with a natural spline and df = 1.

Poisson, Quasi-Poisson and Negative Binomial (NB) distributions for the models were tested and compared in terms of explained Deviance. Since Poisson and NB models showed higher explainability for all causes (Supplementary Figure S2A), the dispersion parameter was analyzed to decide which of the two would be used for each cause (Supplementary Figure S2B). Finally, models using different exposure metrics were compared through AIC metric. All models used 10 lag days, with a natural spline function for the lag component with df = 4. To generate predictions for the crossbasis terms, the mean value was used as the centering (reference) for T_med_ and HI_med_ models. For the remaining models, centering values = 0 were used. An exploratory analysis was conducted to decide whether the day of the week (dow) should be included in the models (Supplementary Figure S3). Since it didn’t appear to have a relevant effect for most causes, we decided not to include it.

### Sensitivity analysis

A sensitivity analysis was conducted for all the possible exposures: T_med_ and HI_med_, amount of hours above a threshold and HAAT metric. For the first two, combinations of natural (“ns”) and B-splines (“bs”) were tested, with *df* ɛ {2, 4, 6} and *df* ɛ {3, 4, 6, 8} for each spline, respectively. For the last two, another set of combinations was considered: natural splines with 1 df, B-splines with 3 df, Cubic regression splines (“cr”) with 3,4,5 and 6 df, and Penalized splines (P-splines) with 5, 10 and 15. Models were compared graphically and through their AIC values.

Relative Risk (RR) estimates regarding each crossbasis term were obtained and used for interpretation of the results. To analyze cut-off points for each exposure metric, we calculated the points where a significant RR of 1.25, 1.5 and 2.0 was crossed, if there were any. For the continuous metrics (T_med_ and HI_med_) the lag effect was also studied in scenarios where

### Software

All analyses were run under the “R” environment, in 4.4.1 version ^52^. Packages “mgcv” (1.9.1) and “dlnm” (v 2.4.7) were used. All the developed code is available on Github: https://github.com/joaohmorais/heat_mortality_RJC/

## Results

During the study period, 390,244 deaths from selected causes and 466,121 deaths from natural causes were registered in RJC. Table 2 describes the summary metrics (median, total and maximum) for death counts due to the 17 cause of death groups included in the study. Some had very low daily median values, especially among the young population. Malignant neoplasms, influenza and pneumonia, and ischemic heart disease had the highest daily median counts for the elderly, excluding the aggregate groups (selected and natural deaths). Some maximum death counts occurred during the Covid-19 critical period (other diseases of the respiratory system, hypertensive disease, undetermined and natural causes), while the maximum for selected causes among the elderly occurred during the November 2023 heat wave (151 deaths in November 18th).

**Table 2.** Summary metrics for cause of death groups included in the study for RJC, Jan/2012-Jun/2024.

| Cause of death group | Daily median deaths |  | Total deaths |  | Max daily deaths (Date) |  |
| --- | --- | --- | --- | --- | --- | --- |
|  | Young | Elderly | Young | Elderly | Young | Elderly |
| Influenza and Pneumonia | 2 | 10 | 10,048 | 46,569 | 10 (Jul 25, 2019) | 32 (Dec 17, 2021) |
| Chronic lower respiratory diseases | 1 | 3 | 3,294 | 14,471 | 7 (Jun 14, 2016) | 13 (Dec 26, 2023) |
| Other diseases of the respiratory system | 1 | 3 | 4,714 | 12,922 | 7 (May 04, 2020) | 14 (Apr 23, 2020) |
| Hypertensive diseases | 1 | 5 | 7,107 | 24,491 | 8 (May 07, 2020) | 20 (Feb 01, 2021) |
| Ischemic heart disease | 4 | 9 | 18,902 | 43,825 | 17 (Aug 05, 2016) | 26 (Aug 17, 2019) |
| Heart failure | 0 | 2 | 3,099 | 10,152 | 5 (Nov 09, 2020) | 10 (Apr 25, 2017) |
| Cerebrovascular diseases | 3 | 7 | 12,596 | 34,807 | 11 (Aug 21, 2022) | 20 (Jun 01, 2012) |
| Other disease of the circulatory system | 3 | 6 | 13,038 | 28,685 | 11 (Oct 11, 2015) | 20 (Jun 14, 2016) |
| Malignant neoplasms | 9 | 16 | 43,832 | 74,805 | 23 (Jun 23, 2016) | 35 (Nov 14, 2016) |
| Alzheimer disease and dementia | 0 | 3 | 276 | 13,157 | 2 (May 24, 2023) | 12 (Nov 24, 2019) |
| Diabetes | 2 | 5 | 8,711 | 22,415 | 9 (Jun 02, 2020) | 15 (Sep 04, 2017) |
| Renal Failure | 0 | 1 | 2,541 | 6,175 | 6 (Mar 11, 2018) | 7 (Feb 05, 2021) |
| Sepsis | 1 | 4 | 4,928 | 19,437 | 7 (Dec 31, 2018) | 15 (Jun 25, 2016) |
| Urinary Tract Disorder | 0 | 4 | 2,121 | 17,361 | 5 (May 29, 2016) | 13 (Feb 08, 2024) |
| Undetermined causes | 3 | 4 | 13,621 | 22,577 | 14 (Feb 02, 2021) | 26 (Jan 30, 2022) |
| Selected causes | 32 | 85 | 148,381 | 390,244 | 69 (Apr 25, 2020) | 151 (Nov 18, 2023) |
| Natural causes | 44 | 99 | 208,996 | 466,121 | 144 (Apr 25, 2020) | 266 (May 02, 2020) |

Table 3 exhibits summary metrics for the climate variables included in the study. Most variables had their highest value during the November 2023 heat wave, when the daily average heat index in the city was 39.69°C. The episode also registered 8 hours of HI >= 44°C and the highest value for HAAT metrics. All metrics depending on the amount of hours of HI above a certain threshold had a value of zero for most of the days in a year, and some even had their 95% quantiles equal to zero as well.

**Table 3.** Minimum, mean/median, 95% and 99% quantiles, and maximum values for climate variables included in the study for RJC, Jan/2012 - Jun/2024.

| Metric |  |  |  |  |  |
| --- | --- | --- | --- | --- | --- |
|  | Min | Mean | Q95 | Q99 | Max (Date) |
| Average daily temperature (°C) | 15.18 | 24.19 | 29.09 | 30.4 | 32.1 (Nov 18, 2023) |
| Average daily heat index (°C) | 14.97 | 25.14 | 31.71 | 33.82 | 39.69 (Nov 18, 2023) |
|  | Avg. Days in year > 0 | Median | Q95 | Q99 | Max (Date) |
| Daily hours of HI ≥ 32°C | 127.68 | 0 | 11 | 15 | 24 (Jan 20, 2024) |
| Daily hours of HI ≥ 33°C | 107.12 | 0 | 10 | 13 | 22 (Jan 20, 2024) |
| Daily hours of HI ≥ 34°C | 88.32 | 0 | 9 | 11 | 19 (Jan 17, 2024) |
| Daily hours of HI ≥ 35°C | 69.44 | 0 | 7 | 10 | 17 (Jan 17, 2024) |
| Daily hours of HI ≥ 36°C | 52.56 | 0 | 6 | 9 | 16 (Jan 17, 2024) |
| Daily hours of HI ≥ 37°C | 38.08 | 0 | 4 | 8 | 14 (Jan 17, 2024) |
| Daily hours of HI ≥ 38°C | 25.28 | 0 | 2 | 6 | 13 (Nov 18, 2023) |
| Daily hours of HI ≥ 39°C | 14.64 | 0 | 0 | 5 | 12 (Nov 18, 2023) |
| Daily hours of HI ≥ 40°C | 8.48 | 0 | 0 | 3 | 12 (Nov 18, 2023) |
| Daily hours of HI ≥ 41°C | 4.1 | 0 | 0 | 1 | 10 (Nov 18, 2023) |
| Daily hours of HI ≥ 42°C | 2.16 | 0 | 0 | 0 | 9 (Nov 18, 2023) |
| Daily hours of HI ≥ 43°C | 1 | 0 | 0 | 0 | 9 (Nov 18, 2023) |
| Daily hours of HI ≥ 44°C | 0.72 | 0 | 0 | 0 | 8 (Nov 18, 2023) |
| HAAT for HI<sub>thresh</sub> = 32°C (°C * hour) | 127.68 | 0 | 43.34 | 70.49 | 185.27 (Nov 18, 2023) |
| HAAT for HI<sub>thresh</sub> = 36°C (°C * hour) | 52.56 | 0 | 9.1 | 25.64 | 117.78 (Nov 18, 2023) |

Expected vs. observed values for daily deaths from selected causes throughout the study period are shown in Figure 1A, along with daily average HI trends (Figure 1B). We can describe the seasonality of deaths with two yearly peaks: a higher one occurring in the fall/winter period (April-June) and the other during the summer period (December, January). This seasonality effect can also be noticed in Figure S1, where the estimated effects for each term in the model is plotted. As deaths due to coronavirus infection (ICD-10 codes B34.2 and U07.1) were not included in the study, the death trend during the Covid-19 period showed a decrease in the death counts. Some notorious excess death periods can be named: in the fall/mid-year of 2016, a mortality burden due to the Chikungunya epidemic in RJC that year was observed ^53^. In 2020 and 2021, three mortality peaks can be pointed: the first in early 2020, after Covid-19 arrival in the country; the second in the period of Gamma (P.1) variant predominance, in early 2021; and the latter during the Influenza A H3N2 outbreak in late 2021 ^47^. Finally, a considerable peak is then observed in November 2023, in two following days: November 18th and 19th - the last two days of November 2023 remarkable heat wave.

**Figure 1.**
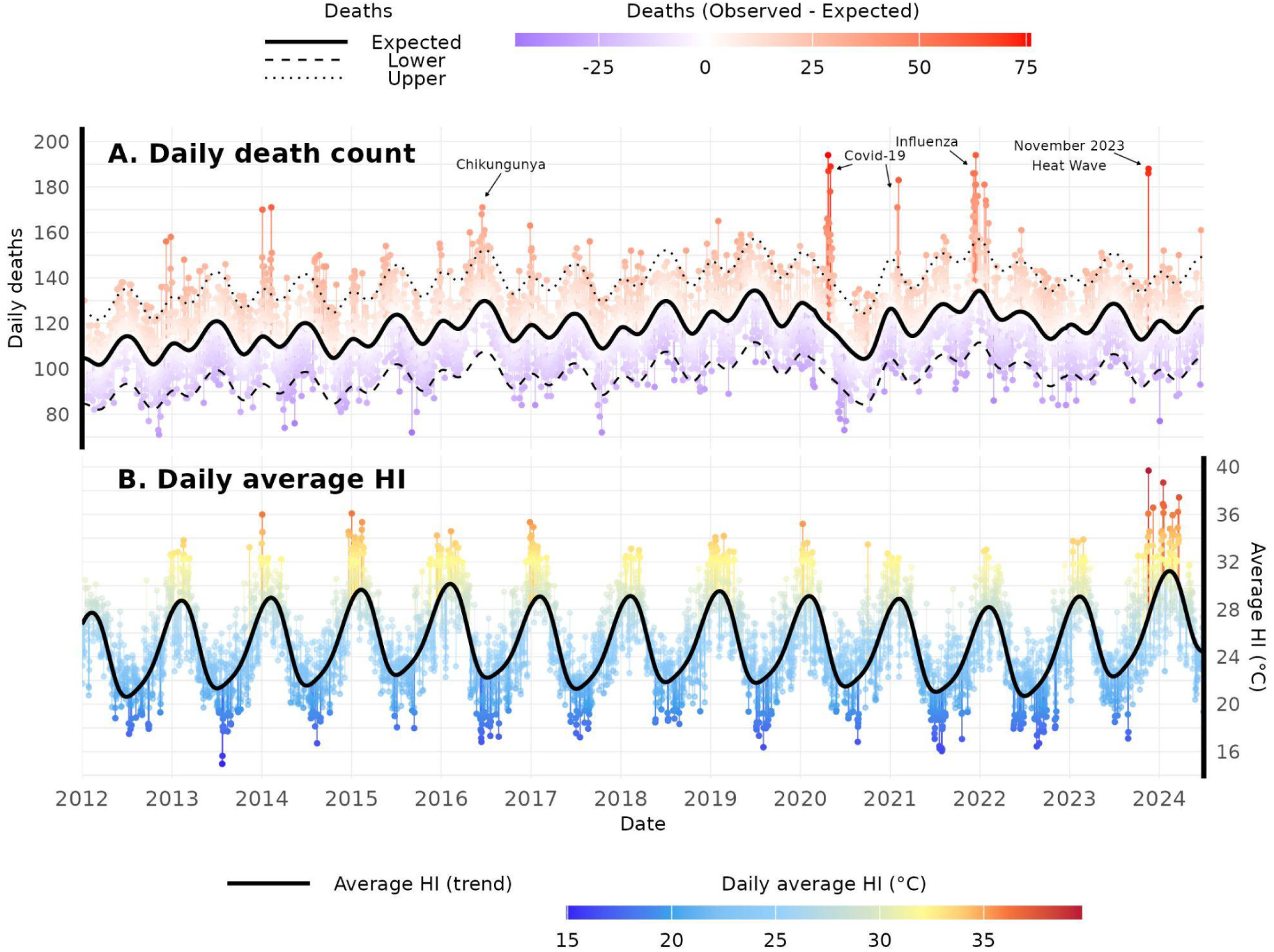
Expected daily deaths (solid line) for all ages in selected causes, and difference between observed and expected (A), and daily average heat index trend (solid line) and observed values (B), for RJC, Jan/2012 - Jun/2024.

The HI yearly patterns expressed in Figure 1B show its expected behavior: an increase during summer months and lower values in the mid-year. Out of the ordinary high values can be seen in the summer season for 2014 and 2015, and 2023/2024 spring and summer. The highest value for average HI registered on November 18th, 2023 was followed by several days with high values as well in the months of December, January, February and March.

### Average daily temperature and heat index

Figure 2 shows the average daily HI (HI_med_) effect for all included causes, among the two age groups. The young (< 65 y.) group presents less cause groups with significant relative risk (RR) in extreme heat conditions. Higher RRs can be observed mainly for undetermined deaths, selected causes, and natural causes; the first two with a broad confidence interval (CI). For the elderly group, on the other hand, most cause of death groups present a significant association between HI_med_ and mortality risk. In these cases, the RR curve seems to get steeper around the 95% quantile of HI_med_, which is 31.71°C (Table 3). The standout groups are Influenza/Pneumonia, hypertensive diseases, ischemic heart disease (IHD), cerebrovascular diseases (CVD), other diseases of circulatory system, Alzheimer/dementia, diabetes, urinary tract infections and undetermined causes. The aggregated groups - selected and natural causes - also show high association. Some of the causes present an increase in RR for low HI_med_ as well, but usually with lower values than those observed for high HI_med_. Heart failure, malignant neoplasms and sepsis show none or little association for both age groups. Similar results for T_med_ are exposed in supplementary Figure S4.

**Figure 2.**
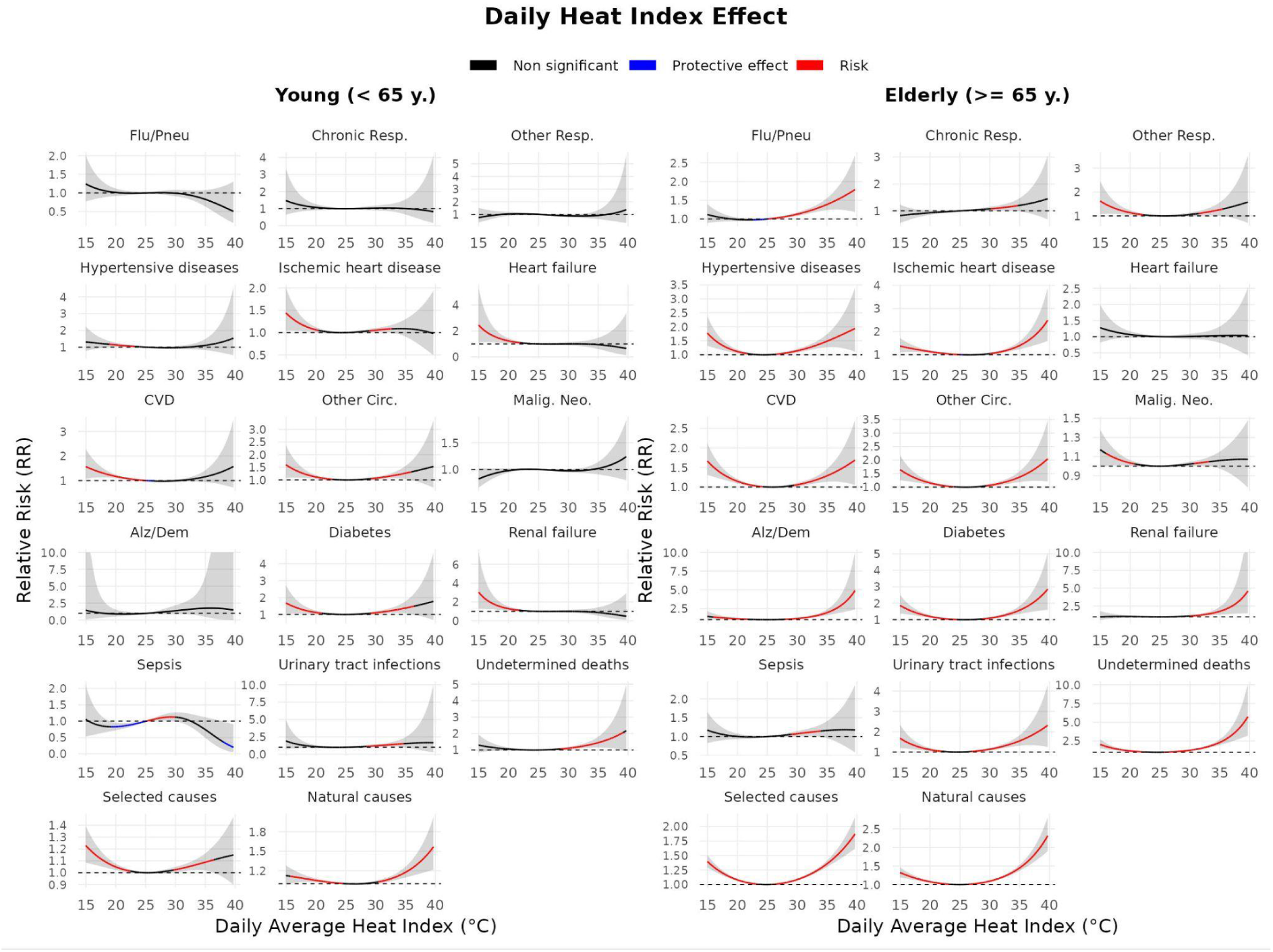
Average Heat Index (HI_med_) effect on mortality for 17 cause of death groups. RJC, Jan/2012 - Jun/2024.

In Table 3, the minimum T_med_ and HI_med_ values necessary to significantly increase mortality risk by 25% (RR > 1.25), 50% (RR > 1.5) and 100% (RR > 2.0) are expressed. For the young age group, a RR of 1.25 is not reached for most causes, except for other diseases of the circulatory system, diabetes, urinary tract disorder, undetermined and natural causes, with the last three reaching RR > 1.5 for some T_med_ or HI_med_ values. The elderly group, on the other hand, showed points of risk increase over at least 25% for all causes but chronic lower respiratory diseases, heart failure, malignant neoplasms and sepsis. A significant RR > 2.0 was observed for 10 out of the 17 causes. The quantile values for T_med_ and HI_med_ necessary to reach such thresholds were similar in general, with slightly lower quantile values for T_med_. It varied from Quantile 87,65% for a RR > 1.25 for deaths of undetermined causes, to Quantile 99.98% to reach a RR of 2.0 in other diseases of circulatory system and selected causes. In the elderly group, on average, a significant RR > 1.25, 1.5, and > 2.0 was reached respectively with quantiles 96.46, 99.11 and 99.82 for HI_med_.

**Table 3.** T_med_ / HI_med_ values (quantiles) necessary to reach significant thresholds of Relative Risks (RR) >= 1.25, >= 1.5 and >= 2.0, for all age groups and included death causes.

| Cause of death group | Young |  |  | Elderly |  |  |
| --- | --- | --- | --- | --- | --- | --- |
| | $RR > 1.25$ | $RR > 1.5$ | $RR > 2.0$ | $RR > 1.25$ | $RR > 1.5$ | $RR > 2.0$ |
| Influenza and Pneumonia | - /<br>- | - /<br>- | - /<br>- | 29.5 (Q96.91) /<br>33 (Q97.94) | 31.6 (Q99.89) /<br>36.9 (Q99.93) | - /<br>- |
| Chronic lower respiratory diseases | - /<br>- | - /<br>- | - /<br>- | - /<br>35.2 (Q99.67) | - /<br>- | - /<br>- |
| Other diseases of the respiratory system | - /<br>- | - /<br>- | - /<br>- | 29.9 (Q98.07) /<br>34.2 (Q99.28) | 31.2 (Q99.76) /<br>36.7 (Q99.91) | - /<br>- |
| Hypertensive diseases | - /<br>- | - /<br>- | - /<br>- | 28.9 (Q94.02) /<br>31.7 (Q94.96) | 30.2 (Q98.69) /<br>34.4 (Q99.45) | 31.8 (Q99.96) /<br>37.7 (Q99.96) |
| Ischemic heart disease | - /<br>- | - /<br>- | - /<br>- | 30.2 (Q98.69) /<br>33.4 (Q98.69) | 31.6 (Q99.89) /<br>35.4 (Q99.74) | - /<br>37.7 (Q99.96) |
| Heart failure | - /<br>- | - /<br>- | - /<br>- | - /<br>- | - /<br>- | - /<br>- |
| Cerebrovascular diseases | - /<br>- | - /<br>- | - /<br>- | 30.1 (Q98.4) /<br>33.9 (Q99.08) | 31.4 (Q99.85) /<br>37.2 (Q99.93) | - /<br>- |
| Other disease of the circulatory system | 30.6 (Q99.21) /<br>34.2 (Q99.28) | - /<br>- | - /<br>- | 30.2 (Q98.69) /<br>33.4 (Q98.69) | 31.8 (Q99.96) /<br>35.9 (Q99.76) | - /<br>38.7 (Q99.98) |
| Malignant neoplasms | - /<br>- | - /<br>- | - /<br>- | - /<br>- | - /<br>- | - /<br>- |
| Alzheimer disease and dementia | - /<br>- | - /<br>- | - /<br>- | 28.5 (Q92.2) /<br>31.2 (Q93.17) | 29.4 (Q96.17) /<br>33 (Q97.94) | 30.4 (Q98.97) /<br>34.9 (Q99.67) |
| Diabetes | 30.1 (Q98.4) /<br>33.2 (Q98.36) | - /<br>- | - /<br>- | 29.2 (Q95.44) /<br>32.5 (Q97.09) | 30.4 (Q98.97) /<br>34.4 (Q99.45) | 31.8 (Q99.96) /<br>36.7 (Q99.91) |
| Renal Failure | - /<br>- | - /<br>- | - /<br>- | 28.7 (Q93.14) /<br>31.7 (Q94.96) | 29.7 (Q97.61) /<br>33.4 (Q98.69) | 30.9 (Q99.52) /<br>35.7 (Q99.74) |
| Sepsis | - /<br>- | - /<br>- | - /<br>- | - /<br>- | - /<br>- | - /<br>- |
| Urinary Tract Disorder | 28 (Q88.74) /<br>30.7 (Q91.57) | 29.2 (Q95.44) /<br>33.7 (Q98.9) | 30.4 (Q98.97) /<br>- | 28.7 (Q93.14) /<br>31.2 (Q93.17) | 29.9 (Q98.07) /<br>33.9 (Q99.08) | 31.2 (Q99.76) /<br>37.7 (Q99.96) |
| Undetermined causes | 29.4 (Q96.17) / 32.7 (Q97.52) | 30.7 (Q99.39) / 34.9 (Q99.67) | 32.1 (Q100) / 37.7 (Q99.96) | 27.8 (Q87.65) / 30 (Q87.93) | 28.9 (Q94.02) / 31.7 (Q94.96) | 29.9 (Q98.07) / 33.9 (Q99.08) |
| Selected causes | - / - | - / - | - / - | 29.7 (Q97.61) / 33.2 (Q98.36) | 31.2 (Q99.76) / 36.2 (Q99.85) | - / 39.4 (Q99.98) |
| Natural causes | 31.9 (Q99.98) / 35.9 (Q99.76) | - / 38.7 (Q99.98) | - / - | 29.5 (Q96.91) / 32.7 (Q97.52) | 30.9 (Q99.52) / 35.2 (Q99.67) | - / 37.9 (Q99.96) |
| **Mean** | 29.4 (Q96.5) / 32.6 (Q97.3) | 29.7 (Q97.42) / 34.6 (Q99.52) | 30.8 (Q99.49) / 37.4 (Q99.96) | 29.2 (Q95.45) / 32.2 (Q96.46) | 30.2 (Q98.63) / 34 (Q99.11) | 30.7 (Q99.37) / 36.1 (Q99.82) |

The lag effects for scenarios when T_med_ and HI_med_ reach the average RR > 1.25 threshold are displayed in Supplementary Figure S5 and S6. Although we see a maximum RR as a result of an immediate (lag 0) effect for most causes, with a decreasing effect as the lag days advance, some causes show a different pattern. Among the elderly, Influenza and Pneumonia show a maximized RR for lags 4-5 for both Tmed and HImed, while Renal Failure and Urinary Tract Infections show a trend of higher RR as the lag days advance.

### Hours of heat exposure

Relative Risks heat-maps for the amount of hours above varying HI thresholds in the elderly age group are shown in Figure 3. A risk gradient is observed for most causes, where mortality risk increases whether the number of hours of exposure or the HI threshold increases. For lower HI levels like HI >= 32°C, higher RRs are observed after a larger exposure period (a 25% increase in risk for natural causes is observed after 13 hours of exposure); for higher thresholds like HI >= 40°C a high RR is observed with fewer hours of exposure (25% increase in risk after 4 hours of exposure). Like the results presented for HI_med_, hypertensive diseases, alzheimer/dementia, diabetes and undetermined deaths are standout groups, while heart failure, malignant neoplasms show little or no association.

**Figure 3.**
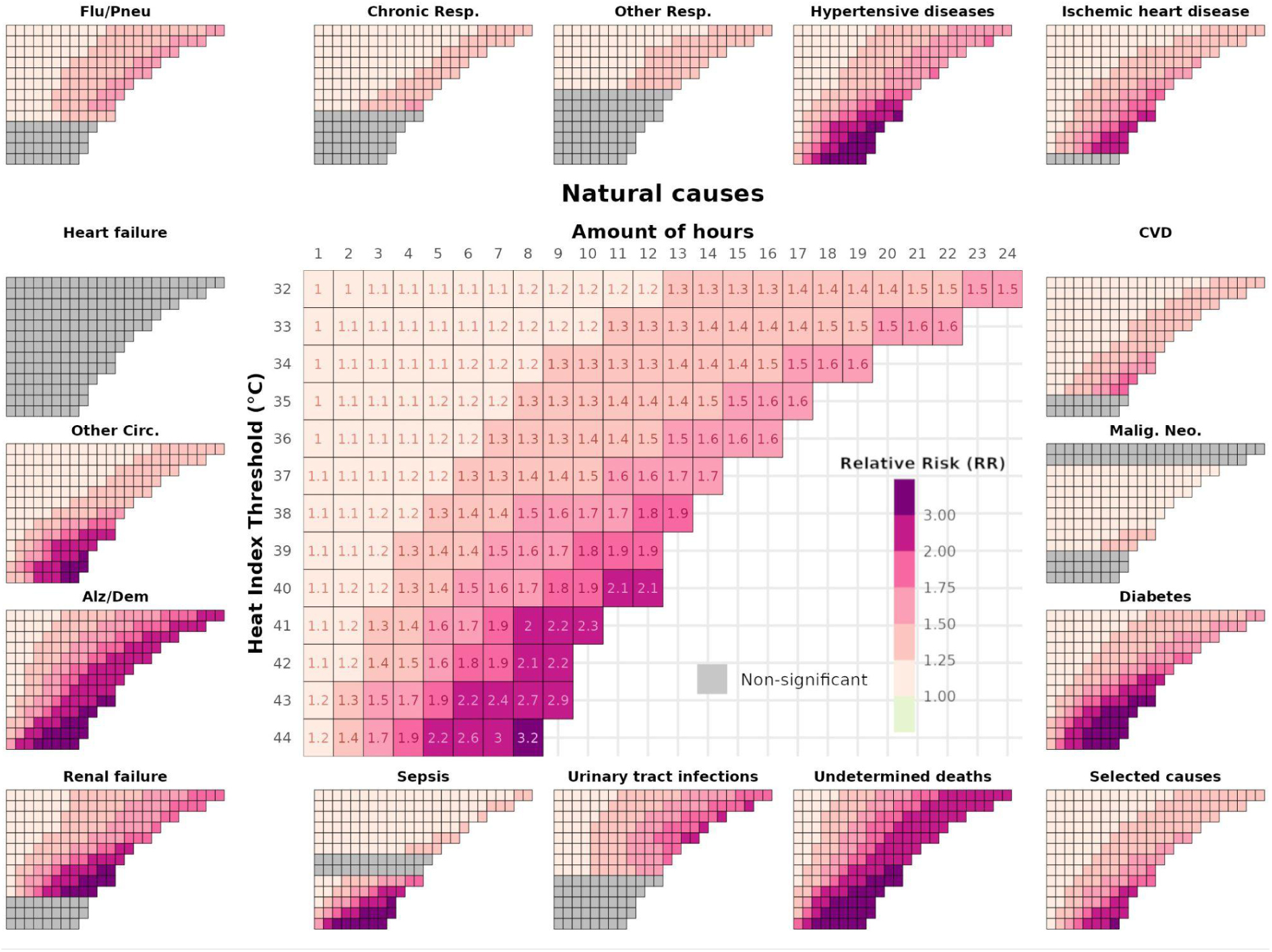
Relative Risks (RR) for amount of hours above a heat index threshold, for 17 cause of death groups among the elderly (>= 65 y.). RJC, Jan/2012-Jun/2024.

Figure 4A illustrates the logic for the HAAT metric calculation. The hourly series for the HI observed during the November 2023 heat wave is represented, with the period of HI above the 36°C threshold filled. The filled area is calculated and represented through the circles on the top. In the November 2023 heat wave, a sequence of extremely warm days was observed, with a scaling number of hours of heat exposure throughout the week, resulting in the highest HAAT value in the whole series, in November 18th (HAAT = 117.78°C*h).

**Figure 4.**
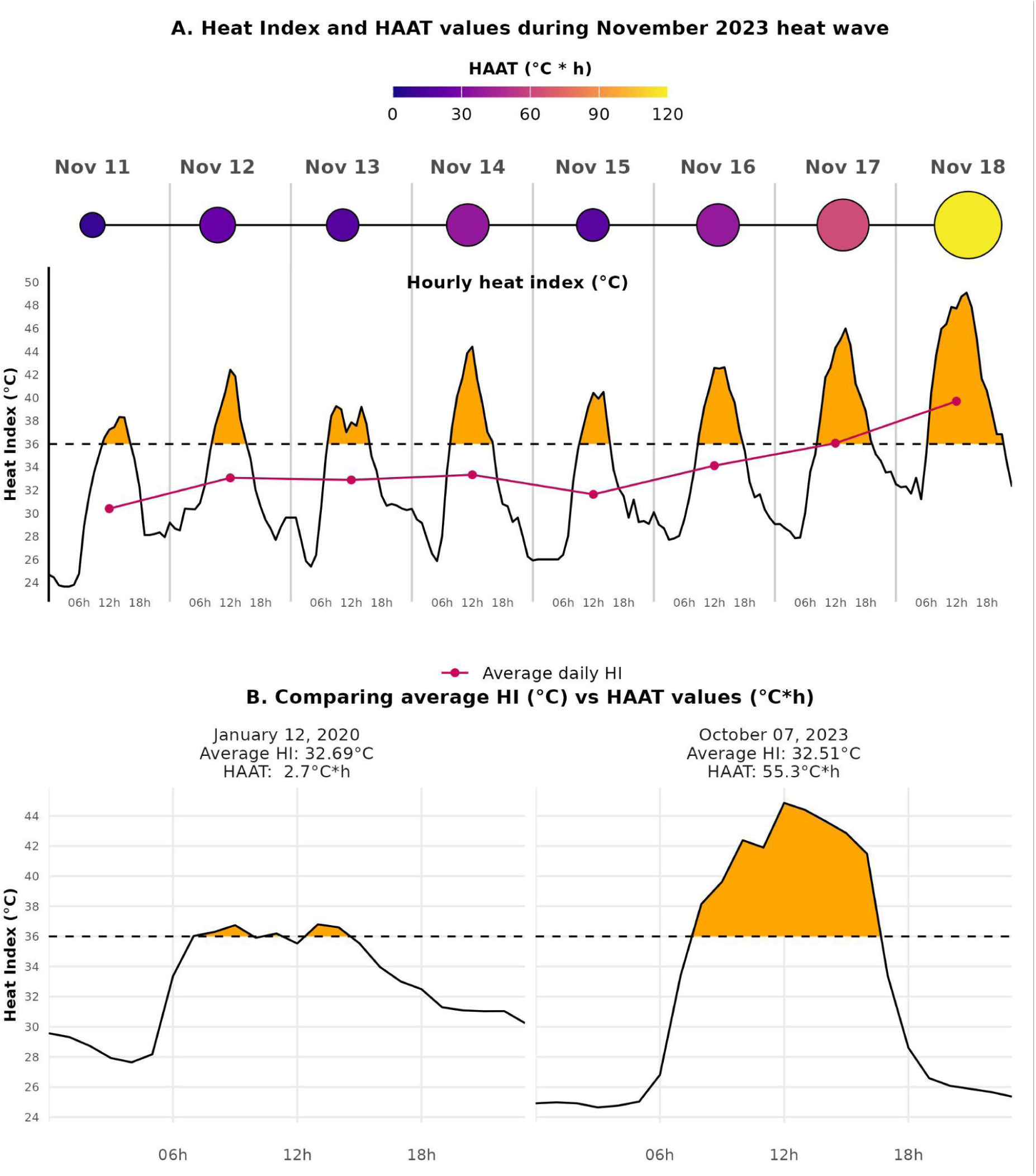
Hourly heat index (HI) values during November 2023 heat wave and respectively Heat Area Above the 36°C Threshold (HAAT) calculated values (A); in (B), the difference in HAAT values between two dates (Jan 12, 2020 and Oct 07, 2023) with similar average HI values.

In Figure 4B two days with similar HI_med_ values are compared, showing considerable differences in their respective HAAT values. Even though Jan 12, 2020 had a higher average HI than October 7h, 2023, the latter had a much higher area above the HI >= 36°C limit. This difference in the exposure period is captured through the HAAT metric and now through averaged daily values.

The AIC metrics for the models using each of the included exposure variables - T_med_, HI_med_, amount of hours above HI threshold, HAAT metric and HW definition (single and added effects) - are compared in Figure 5. Since several different models were fitted for each of the last three groups mentioned, only the one with lower AIC was considered for that group. Models using the HAAT metric performed best for 10 out of the 17 causes (mostly, when used along with T_med_ - 8/10). Amount of hours performed best among the groups with low death counts; T_med_-only was best for 2 groups and HW models (either single or added effects) for other 2. HAAT showed, in general, better performance when included along with T_med_ in the models. Complete list of results are reported in the Supplementary Material.

**Figure 5.**
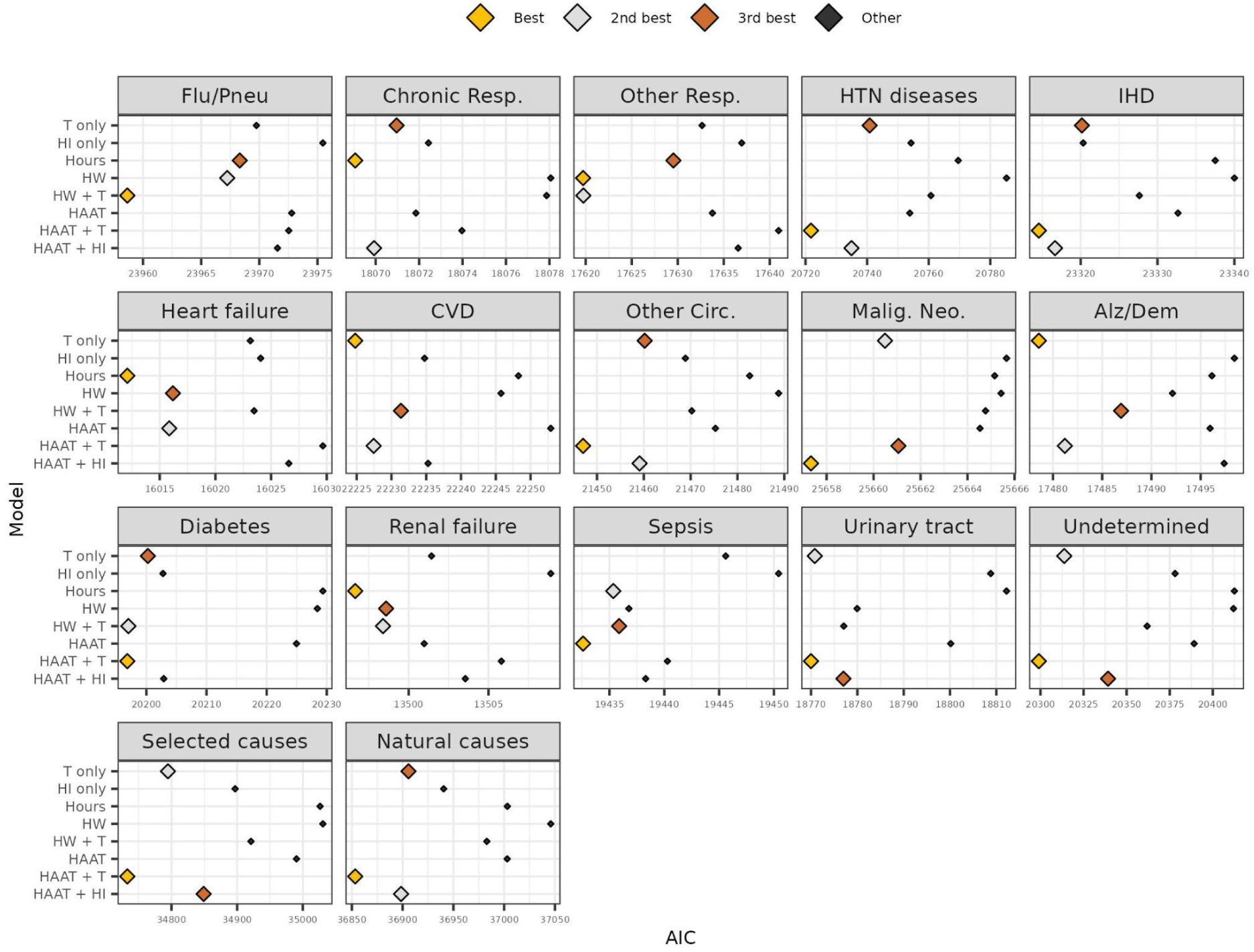
AIC values for the best model among different exposure variables: average temperature (T only), average heat index (HI only), amount of hours above a heat index threshold (Hours), heat wave definition (HW), heat wave definition + average temperature (added effects, HW+T), HAAT values, HAAT values + average temperature (HAAT + T), HAAT values + heat index (HAAT + HI) for 17 cause of death groups in the elderly. RJC, Jan/2012-Jun/2024.

Supplementary Figure S7 compares whether using accumulated HAAT helps explain mortality any better than single-day HAAT, and which HI_thresh_ performs best. When not used along with an average metric (T_med_ or HI_med_), a HAAT with HI_thresh_ = 32°C shows better explainability - which makes sense, since it has fewer days with null values than HI_thresh_ = 36°C (Table 3). However, in most scenarios the use of HAAT along with T_med_ showed a better performance - and in that case, the use of single-day HAAT calculated with a HI_thresh_ = 36°C was a best-performing combination.

Figure 6 exposes the relationship between the defined thresholds in RJC’s protocol and the mortality risks for diabetes, hypertensive disease, IHD, renal failure and natural and selected causes. Since the triggers for the protocol are based on the amount of hours above the thresholds of HI >= 36°C, HI >= 40°C and HI >= 44°C, the mortality risks are expressed in terms of these three variables. They are compared with the risk curves obtained using the HAAT metric instead, calculated with a HI threshold of 36°C. The model results indicate a 25% increase in mortality risks when thresholds of 6 hours or higher with HI >= 36°C are reached (min.: 5.5 hours for Renal Failure; max.: 8.4 hours for selected causes). An increase of 50% is only reached with an amount of hours with HI >= 36°C higher than 10 (min.; 9.9 for Renal Failure; max.: 15.3 for selected causes). When looking at the amount of hours with HI of 40°C or higher, these marks are crossed much quicker. 1.8 and 3.9 hours are enough to reach a 25% increase for Renal Failure and selected causes respectively; 50% is reached with 3.3 and 7.1 hours. A doubled risk of mortality is reached for all causes shown expected for the selected causes, when the limit of 5.7 hours with HI >= 40°C (renal failure) or 11.3 hours with HI >= 40°C (IHD). Even shorter periods of exposure are necessary when looking at the hours with HI above 44°C curve. 1 hour is needed to increase the mortality risk for hypertensive diseases and renal failure, while 5 hours already doubles the risk for selected causes. Similar, but simpler, trends are observed when analyzing the heat area (HAAT) effect: lower thresholds are necessary for renal failure mortality risk to increase (28 °C*h for a 50% increase, 48 °C*h to double), while for selected and natural causes this increase in risk is observed much later (69.6°C*h for a 50% increase, 93 °C*h to double for selected causes). A similar visualization for all causes included is shown in Figure S8. From that visualization, we see that, across all causes, the median HAAT value where risk of mortality among the elderly increases by 25% is 18.44 °C*h; it increases by 50% with HAAT >= 48.77°C*h; and doubles with HAAT >= 69°C*h.

**Figure 6.**
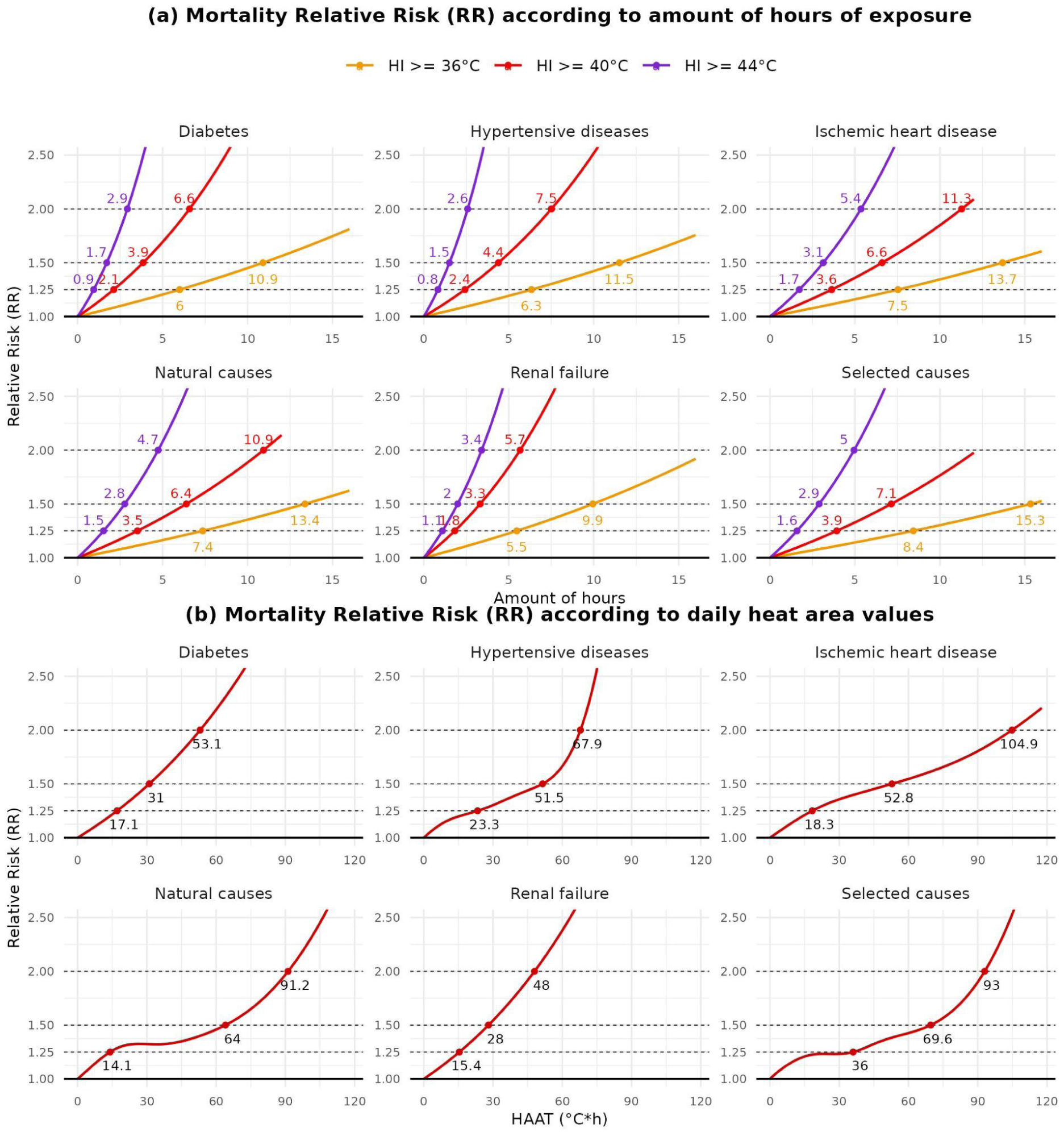
Relative Risk thresholds (25%, 50% and 100% of excess mortality) according to amount of hours of exposure (a) and according to accumulated heat area values (b) for the elderly group. Labeled points represent points of crossing established risk marks (1.25, 1.5, 2). RJC, Jan/2012-Jun/2024.

## Discussion

This study evaluated the effect of heat exposure in mortality due to 17 causes and among two age groups, in Rio de Janeiro city from January 2012 to June 2024. The studies with similar purposes found did not consider these amount of groups, nor covered the most recent period of important heat wave episodes in Brazil (November 2023 and January 2024). It also compares different metrics of heat exposure in terms of amount of exposure time: we start by introducing the amount of hours above a HI threshold to the models, and then present the HAAT metric.

### Death causes and age groups

We explored how continuous T_med_ and HI_med_ relate to mortality. Even though patterns for both metrics were similar when compared, the punctual risk estimates for extreme values of HI_med_ were higher and had larger confidence intervals (CIs). This can be justified by the fact that amplitude for HI_med_ was higher - it had lower minimum and higher maximum values than T_med_ (Table 3). While this may help on the discrimination of days on extreme heat (moist heat, specifically) situations, it also increases uncertainty on the effect estimates since there are less observations concentrated in the tails of the variable distribution.

As expected, fewer causes of death showed significant heat-mortality associations for the young group, when compared to the elderly (Figure 2). Apart from the aggregate causes (selected and natural), only other circulatory diseases, diabetes, urinary tract infections and undetermined deaths presented a significant increase in RR for mortality in the young as Tmed got higher (Figure S4). Both aggregate causes showed a similar pattern for mortality, but with RRs much lower than those obtained for the elderly. These findings are consistent with similar studies that show age as a considerable determinant in the heat-mortality relationship ^15,33,54^.

There was a significant heat-mortality association for almost all causes in the elderly group. Chronic lower respiratory diseases, other respiratory diseases, heart failure, malignant neoplasms and sepsis were the 5 groups that showed no significant risk for high HI_med_ values. The other respiratory diseases group did show, on the other hand, a high significant RR for high values of T_med_. This relates to the discussion on the complexity of the role humidity plays in the heat-mortality relationship ^50,51^. Low H and high T episodes (on which HI would be lower than T) can lead to respiratory issues ^51,55^; additionally, so can pollution and bad air quality ^54,56,57^, which were not considered in this study. The humidity discussion will be prolonged later on. Heart failure and sepsis are groups that besides having low death counts (2nd and 4th lowest median daily counts among the included groups, Table 2), are also unspecific and usually not the underlying cause of death reported (variable upon which the study is based on). The ongoing recommendation by the Brazilian Ministry of Health is not to use non-specific ICDs such as heart and respiratory failure or sepsis - so called “garbage codes” - as underlying cause of death ^58^. Finally, even though higher mortality due to neoplasms during heat waves has been reported in the literature ^59–61^ and majorly attributed to harvesting effect ^60^, we have not seen a significant effect in this study.

The heat-mortality relationship observed for IHD, CVD and other circulatory diseases in the elderly are consistent with the literature. The pathophysiological mechanisms in this relationship have been extensively described ^11,22,62,63^ and involve a diminished capacity of heat stress responses, leading to higher cardiac demand and strain on the cardiovascular system. A recent meta-analysis including studies from more than 20 countries showed a 2.1% increase in cardiovascular mortality for every 1°C increase in temperature, with cause-specific analyses showing higher risks for stroke, coronary heart diseases and heart failure ^62^. A cardiovascular effect was also seen in a study involving 326 Latin American cities ^15^; and in studies that took place in Brazilian cities ^4,23,30,64^.

Deaths due to diabetes also had a strong association with heat exposure - a 25% increase in days with HI_med_ >= 32.5 and 100% increase for HI_med_ >= 36.7 (Table 3) for the elderly. Even though the relation of heat exposure with diabetes is still not totally clear, it is argued that diabetic patients have lower skin blood flow and thermoregulation mechanisms, which affect blood sugar control and cardiovascular regulation ^65^. Song *et al.* recent review ^65^ of 18 studies shows a significant heat-mortality relation for diabetes, with a higher impact on mortality than morbidity. The strong association found in this study is supported by previous studies in Brazil: In a 2020 study in Rio de Janeiro Metropolitan Region, Geirinhas *et al.* found that the highest excess mortality identified was due to diabetes, and particularly in women and the elderly ^32^. In addition to it, Zhao found a high association between heat wave episodes and hospitalization due to endocrine, nutritional and metabolic diseases in Brazil during 2000-2015 ^17^.

Mortality by Alzheimer and dementia was also strongly associated with extreme values of T_med_ and HI_med_ among the elderly. The association showed even lower thresholds necessary to reach a 25% increased risk of death - a T_med_ of 28.5 (92.2% quantile) or a HI_med_ of 31.2 (93.17% quantile). Alzheimer’s disease admissions had already been shown to be related to heatwave days ^66^. Xu *et al.*, in a retrospective cohort study in Australia, reported an increased risk of hospitalizations due to Alzheimer’s disease during heatwaves, and reviewed plausible mechanisms, which include: a change in thermoregulation capacity due to the use of neuroleptics; a dopamine deficit during heatwave days; and a mixed capacity of recognizing hostile environments on extreme temperatures ^67^. Other two groups also showed low thresholds for a mortality increase among the elderly: renal failure and urinary tract disorders. Heat effects over urologic outcomes - which include kidney disease, renal failure, urolithiasis and urinary tract infection - have been documented in studies occurring in Australia ^18,68^ and in American cities ^69,70^. In Brazil, Zhao *et al.* ^17^ also reported strong associations between severe heatwaves and admissions by genito-urinary causes. Heat exposure raises the potential of dehydration, which is already critical in the elderly ^71^. This reduces blood volume and urine output, which can impair renal hypoperfusion and lead to kidney injury, or increase the risk of urinary tract infections.

Undetermined and aggregated causes present evident risk increases when high values of T_med_ or HI_med_ are reached, for both age groups (except for selected causes among the young). Undetermined deaths showed higher values of RR, followed by Natural causes. This suggests that there can be an excess use of garbage codes specifically in periods when there is a mortality excess during heat exposure days, which needs a deeper look.

Although not the focus of the study, high RRs were also observed for periods of colder temperatures for both metrics. The effects were, in most cases, lower than those observed on warm days, and those findings are also consistent with the literature ^23,30,56,72^.

### Exposure period to heat as a metric for explaining heat-related mortality

We were interested in investigating how the amount of time individuals stay exposed to high heat thresholds affect mortality risk. Since this exposure time is not exactly captured through daily average metrics (Figure 4B), we did that from two perspectives: analyzing the amount of hours spent above varying HI thresholds on each day; and through the HAAT metric. While these metrics carry less explanatory potential - once they have zero values for most days (Table 3), their inclusion in the models for mortality can help distinguish days of unusual heat exposure and aid the definition of thresholds for HWSs.

The models analyzing hourly exposure to HI thresholds revealed a gradient effect for most causes of death in the elderly group (Figure 3), with increased risk associated with both longer exposure periods and higher HI thresholds. The gradient effect is more pronounced for the aggregated causes, undetermined deaths, circulatory causes, Alzheimer/dementia and diabetes. As reported previously, heart failure and malignant neoplasms showed none or few scenarios of increased risk. For respiratory causes, models for higher HI thresholds showed no significance either. This again can relate to the discussion on the complex role humidity plays in population mortality studies, as well as other factors that influence respiratory mortality that were not included.

From both Figures 3 and 6 we see that, for lower HI thresholds, higher mortality risks are reached with prolonged exposure period; in contrast, shorter exposure times are needed for higher HI thresholds. Figure 6A exhibits that a 7.4 hours and a 13.4 hours exposure period to HI above 36°C in a given day are associated with a 25% and 50% increase in the mortality risk by natural causes, respectively. For a HI >= 40°C threshold, the exposure periods for reaching the same marks reduce to 3.5 and 6.4 hours, and 1.5 and 2.8 hours are needed for HI >= 44°C. Associations between HAAT values and mortality risk show similar patterns (Figure 6B), and are simpler to interpret since they do not depend on different HI thresholds. Figure S8 shows that, across all causes for the elderly, a HAAT of 18.44°C*h is the median value leading to a 25% risk increase; 48.77°C*h leads to a 50%, and 69°C*h leads to a 100% increment.

Differently from other metrics used on heat-mortality studies or HWSs, the HAAT metric is based on the amount of exposure time to high heat (HI >= HI_thresh_) in a given day (or days), rather than summarizing heat exposure in an average daily metric. It is, however, not exclusive to the Heat Index, since its logic can be used to any other hourly metric or even temperature. Its explanatory potential was compared to the other metrics included in the study (Figure 5). For most causes (13 out of 17), HAAT composed variables (either HAAT-only, or added to HI_med_ or T_med_) performed better than the commonly used combination of a heat wave indicator + T_med_, including for the broadest groups of selected and natural causes. This shows that the proposed HAAT metric succeeds on capturing days with unusually large exposure to high heat, and shows relevant explanatory potential for explaining heat-related mortality, specially when used along with a summary metric (like T_med_).

### RJC’s heat Protocol

This study offers evidence to support RJC’s heat protocol implemented in June/2024. Through Figure 6 we can investigate whether the defined cut-off points in the protocol - 4 hours of HI >= 36°C (levels NC2 and NC3), 4 hours of HI >= 40°C (level NC4) and 2 hours of HI >= 44°C (level NC5) - were appropriately established. The first cut-off point, 4 hours of HI >= 36°C, does not show relevant increases in mortality by itself, since an increase in mortality by 25% is only seen after around 6 hours of exposure (5.5 hours for Renal Failure, 6 for Diabetes). The 4 hours of HI >= 40°C threshold defined for level NC4 represents higher risk levels - a 50% increase in mortality by diabetes (3.9 hours), renal failure (3.3 hours), and HTN diseases (4.4 hours) and a 25% increase in IHD, natural causes and selected causes. Finally, the 2 hours of HI >= 44°C trigger represents more than 50% increased mortality for diabetes, HTN diseases and renal failure.

Even though the first threshold in the protocol does not lead to a significant increase in mortality, it doesn’t mean it is overestimated. The second and third levels (NC2 and NC3) in the protocol involve measures of “monitoring, alert and communication” to the population ^40^; while response from the health system and adaptation of public activities are only considered in level four (NC4), when an increase in mortality is evident. In addition to it, a mortality excess is an extreme outcome - before that, an increase in hospital attendances and morbidity may be observed, as documented in other studies ^17,73–75^. Analyses of these more proximal outcomes would be great evidence to better define a heat-related risk scale. As on the upper levels of the protocol, there seems to be an agreement as well: While 2 hours of HI >= 44°C (fifth level, NC5) do not lead to the maximum RR mark considered (RR >= 2.0), one more hour of exposure would reach such mark on HTN diseases (2.6 hours), renal failure (3.4 hours) and diabetes (2.9 hours).

RJC joins an extensive list of cities that have a HWS implemented, though very few are Latin American cities ^37,76^. It considers the measurement of temperature along with humidity through the calculation of the HI, as it is done in Switzerland ^77^ and by the National Weather Service (NWS) in the United States ^78^. Even though the index is a commonly used metric to express thermal comfort ^41,79^, it was never designed to explain heat-related mortality^51^. This study shows that using HI can help distinguish very unusually warm days (moist heat, mostly) but HI doesn’t seem to explain mortality any better than temperature (Figure 5), as reported in studies using composite indices ^50,51^. The duration of the heat event is also taken into consideration, since a single warm day can’t go any further than level 2 (NC2) ^40^, following common agreements on heat wave definitions ^80^. Despite using HI as its basic metric, it differs from other existing HWSs by considering the amount of exposure necessary to trigger a new level, rather than reaching a specific daily average/maximum T or HI value ^37^, since days with similar averaged values may have much different exposure times (Figure 4). This study offers, finally, the possibility to put together the exposure period along with HI thresholds through the HAAT metric. Simpler cut-off levels could be defined based on the metric breaks leading to increased mortality risk - as on natural causes of death, or the median values across all causes (Figure S8).

### Limitations and future work

This study prioritized studying the association of heat exposure with specific mortality causes in the most recent years in Rio de Janeiro. Disaggregating by cause groups led to low death counts, which made difficult the consideration of broader age groups, sex, race or socio/spatial differences in the analyses, even though evidence shows they play a significant role in the heat-related mortality ^4,24,25,81^. Pregnancy and maternal outcomes are also stated as related to heat exposure in the literature ^20,21^, and must be investigated in further studies for the RJC population.

Spatial differences in heat exposure and its related outcomes were overlooked. RJC is a large, urban city, prone to the urban heat island (UHI) effect ^82^, where densely populated and urban areas, as well as those with lower vegetation cover, tend to accumulate more heat^83^. These areas are also highly correlated with unfavorable socio-economic conditions ^26,84^. Moreover, gaps in weather stations data also impose a challenge: many densely populated areas of the city lack station coverage (Figure S9), while part of the stations are located in airports or coastal locations, which do not capture intra-urban heat heterogeneities ^51^.

The model structure must be brought to discussion as well: the inclusion of a smooth term to account for the differential patterns in mortality during Covid-19 pandemic more acute period can mask relevant patterns when examining heat-related mortality. Sousa *et al.* ^85^ study in Portugal showed that mortality excess waves during the 2020 summer were not solely attributed to Covid-19; rather, they suggest that heat-related mortality may have been exacerbated during the lockdown period, possibly due to patients not seeking medical attention or being unable to be admitted to emergency rooms due to concurrent capacity issues during the pandemic. This effect, if similar in RJC, would not be captured by the crossbasis terms in the models and can be further investigated.

Final considerations on study limitations regard the complex role humidity plays on heat-related mortality. Even though it is clear that humidity strongly affects heat-related outcomes in human physiological studies ^10,86^, this effect is not clear in population studies ^51^. Moist heat affects individuals’ capacity to dissipate heat and therefore to cool themselves. This effect, when present for a large exposure time, can lead to heat exhaustion or heat stroke, particularly among the most vulnerable ^25,87^. However, epidemiological studies conducted considering humidity along with temperature ^50^ or through a composite index ^88–91^, have shown little or no impact on explaining mortality. Our findings contribute to this trend, since models for HI_med_ had similar or higher AIC than T_med_ in all the cause groups studies (Figure 5). Baldwin *et al*. ^51^ contributes by pointing out the fragility of using such composite indices, since increases can be observed due to elevations in temperature, in humidity, or in both. Including both temperature and humidity (preferably, not relative humidity) in the mortality models - possibly, with an interaction or effect modification term - is encouraged and could help better understand how T and H relate to each of the mortality causes.

### Final considerations

Exposure to high heat levels for prolonged times lead to an increase in mortality in Rio de Janeiro, affecting mainly the elderly population. The young are affected in smaller proportion and in a fewer number of causes, while Diabetes, Alzheimer/Dementia, Renal Failure and Hypertensive diseases are standout groups among the elderly. Modeling the amount of hours exposed to high heat index extremes showed the thresholds defined in RJC’s protocol are adequate, though adjustments can be made to match defined 25%, 50% and 100% increase in risk marks. The approaches taken in the study suggest that the consideration of the exposure time to high heat thresholds, either through the amount of hours of exposure or through the HAAT metric, is relevant and can help explain mortality in unusually warm days, as well as guiding definition of cut-off points for Heat Warning Systems.

## Supporting information

Supplementary Material: Model Result List

## Ethical statement

The authors inform that the climate data used in the study are public and freely available for analysis. The mortality data, however, is not public and is part of a project approved under opinion no. 6.572.784 by the Research Ethics Committee of the Municipal Health Department of Rio de Janeiro. No personally sensitive or identifiable information was used in the analysis.

## Authors contributions

All authors participated in the conception and design of the work. JHAM, VS and OGC worked on statistical modeling and data analysis. All authors participated in the interpretation of the results. All authors participated in the manuscript writing and its critical review. All authors worked on the approval of the final version of the manuscript.

## Data availability

Climate data and R code used are publicly available in the github page https://github.com/joaohmorais/heat_mortality_MRJ_article. As mentioned in the ethical aspects section, mortality data cannot be provided.

## Declaration of conflicts of interest

The authors declare they have no conflicts of interest related to this work to disclose.

## Acknowledgments

We would like to thank Rio’s Municipal Health Secretariat (SMS), Operations and Resilience Center (COR), Alerta Rio, and Municipal Secretariat for Public Services and the Environment (SMAC) for collaborative work.

## Notes

### Competing Interest Statement

The authors have declared no competing interest.

### Funding Statement

This study did not receive any funding.

### Author Declarations

The Research Ethics Committee of the Municipal Health Department of Rio de Janeiro approved this study under opinion no. 6.572.784.

