## Supplementary Material for "Quantifying heat exposure and its related mortality in Rio de Janeiro City: evidence to support Rio’s recent heat protocol"

**Figure S1. GAM model terms effect: (a) f1, the long-term trend effect over *t* (date of death); (b) f2, the cyclic day-of-year (doy) effect; (c) f3, the differential death pattern during Covid-19 pandemic.**

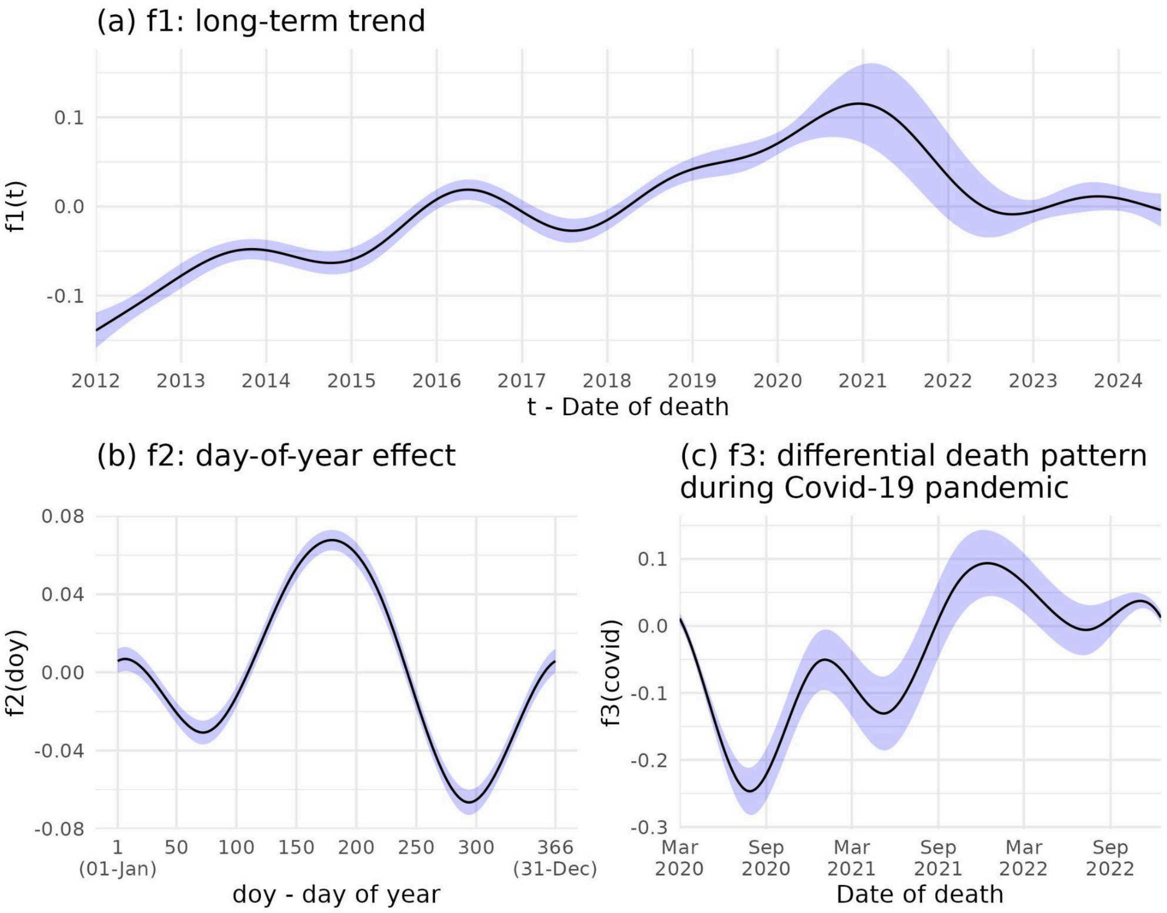


**Figure S2. Explained deviance for mortality models by each cause among the elderly, varying model likelihood: Poisson, Negative Binomial and Quasipoisson (A). In (B), the calculated dispersion parameter (Deviance/Residual df) for Poisson models.**

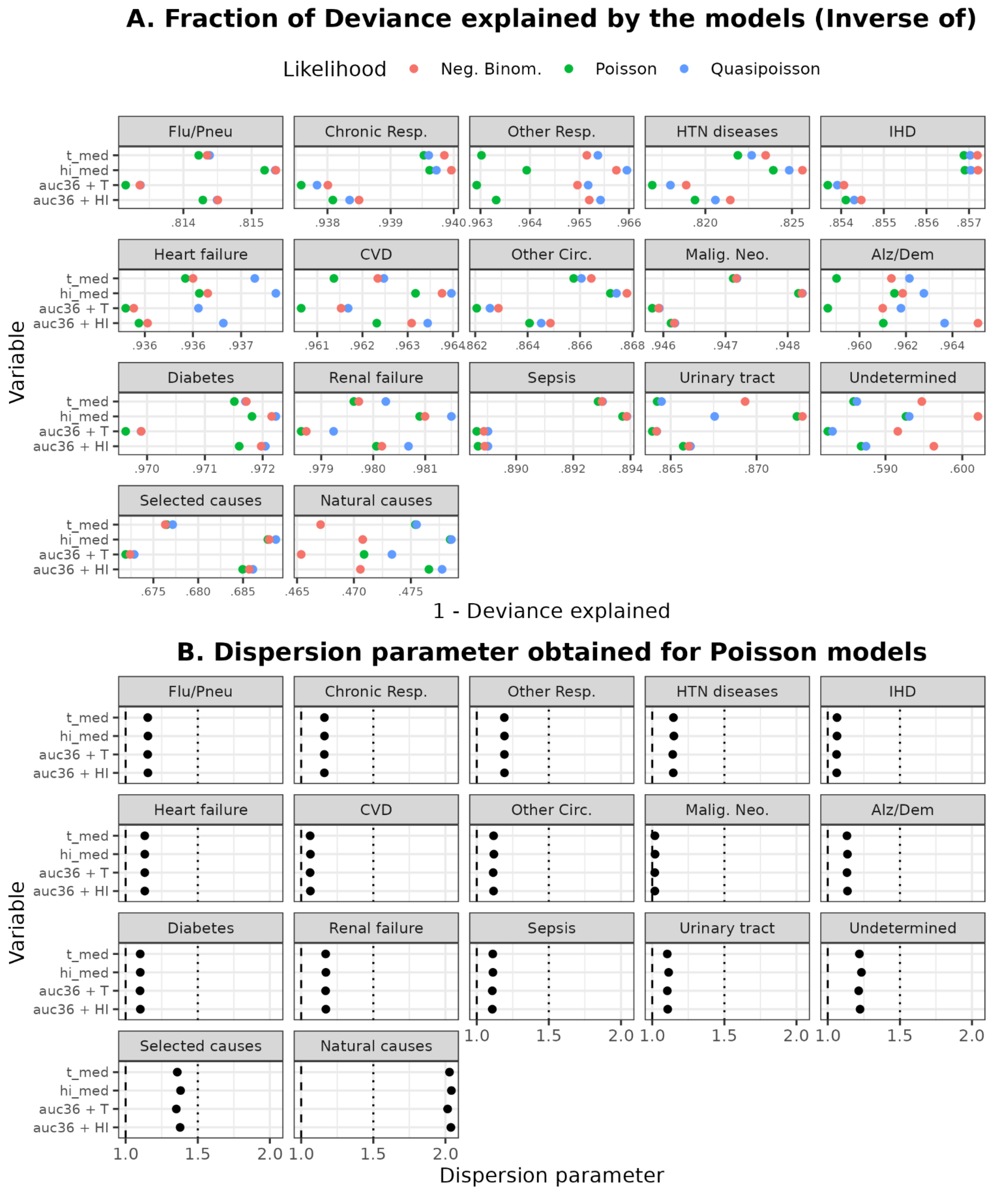


**Figure S3. Death count distribution by day of the week, for each cause included, among the elderly (A) and p-value obtained for simple GLM models using day of week as a covariate (B).**

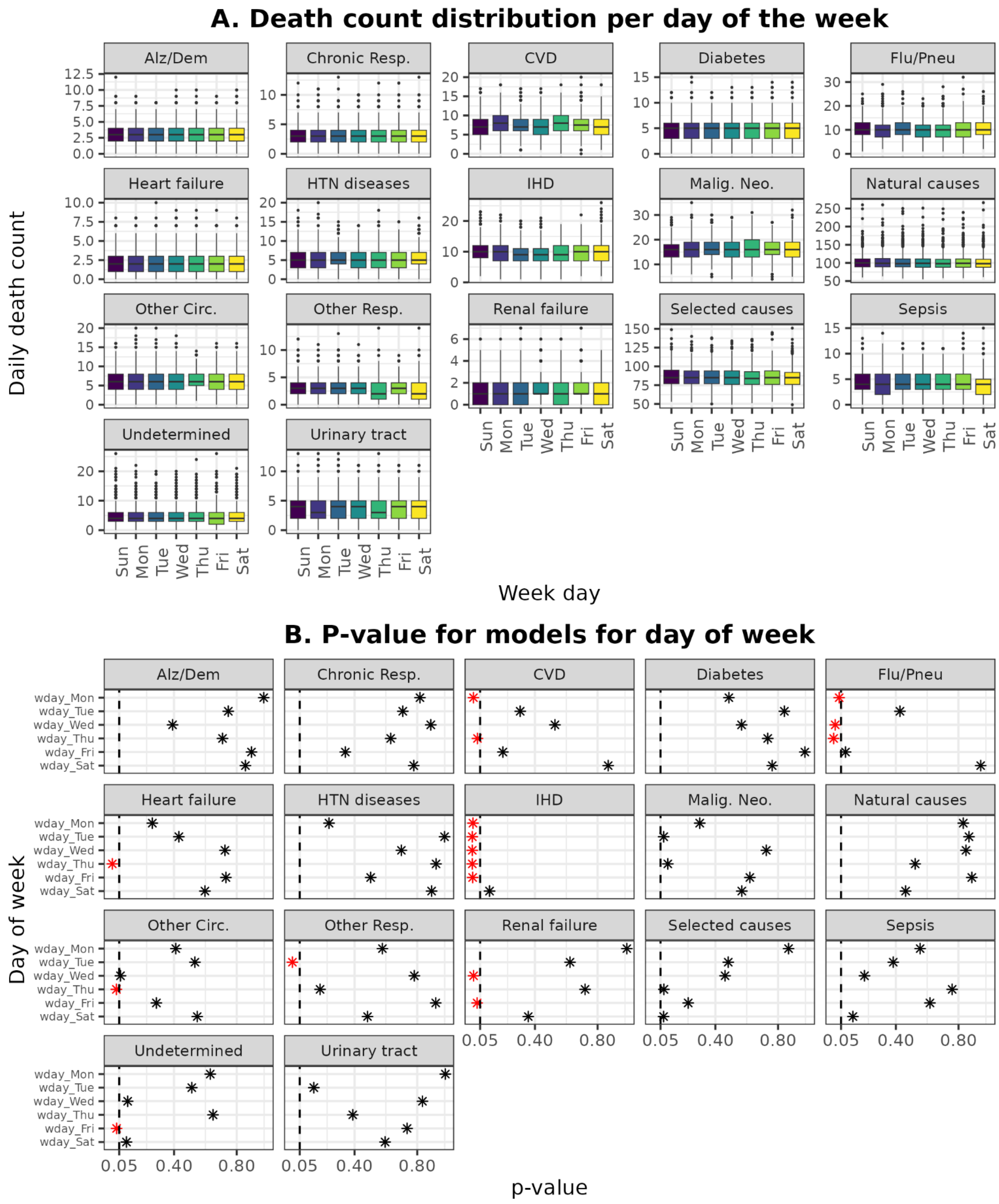


**Figure S4. Average Temperature ($T_{med}$) effect on mortality for 17 cause of death groups. RJC, Jan/2012 - Jun/2024.**

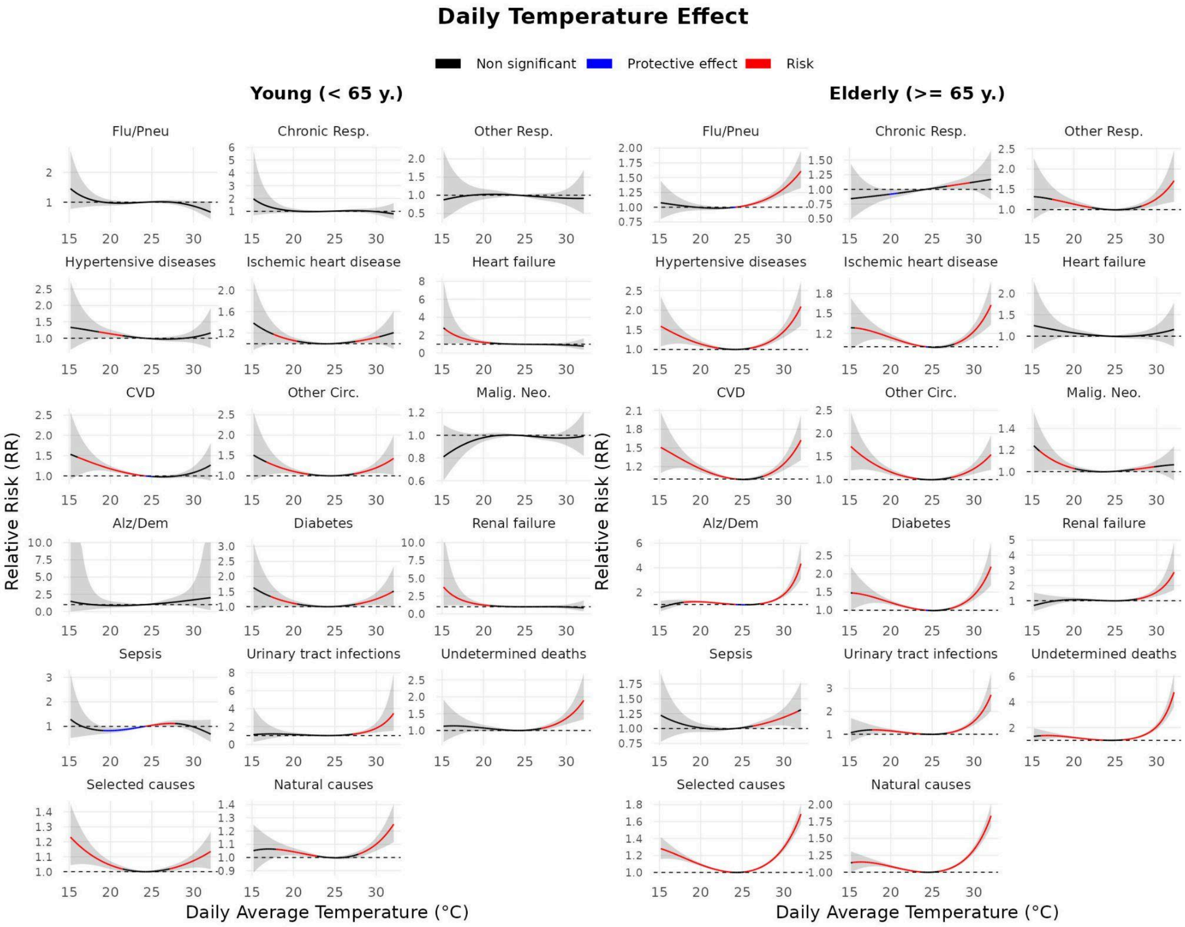


**Figure S5. Mortality Relative Risk (RR) when exposed to high HImed (RR > 1.25 threshold) according to lag days, for all causes included among young (left) and elderly (right). RJC, Jan/2012-Jun/2024.**

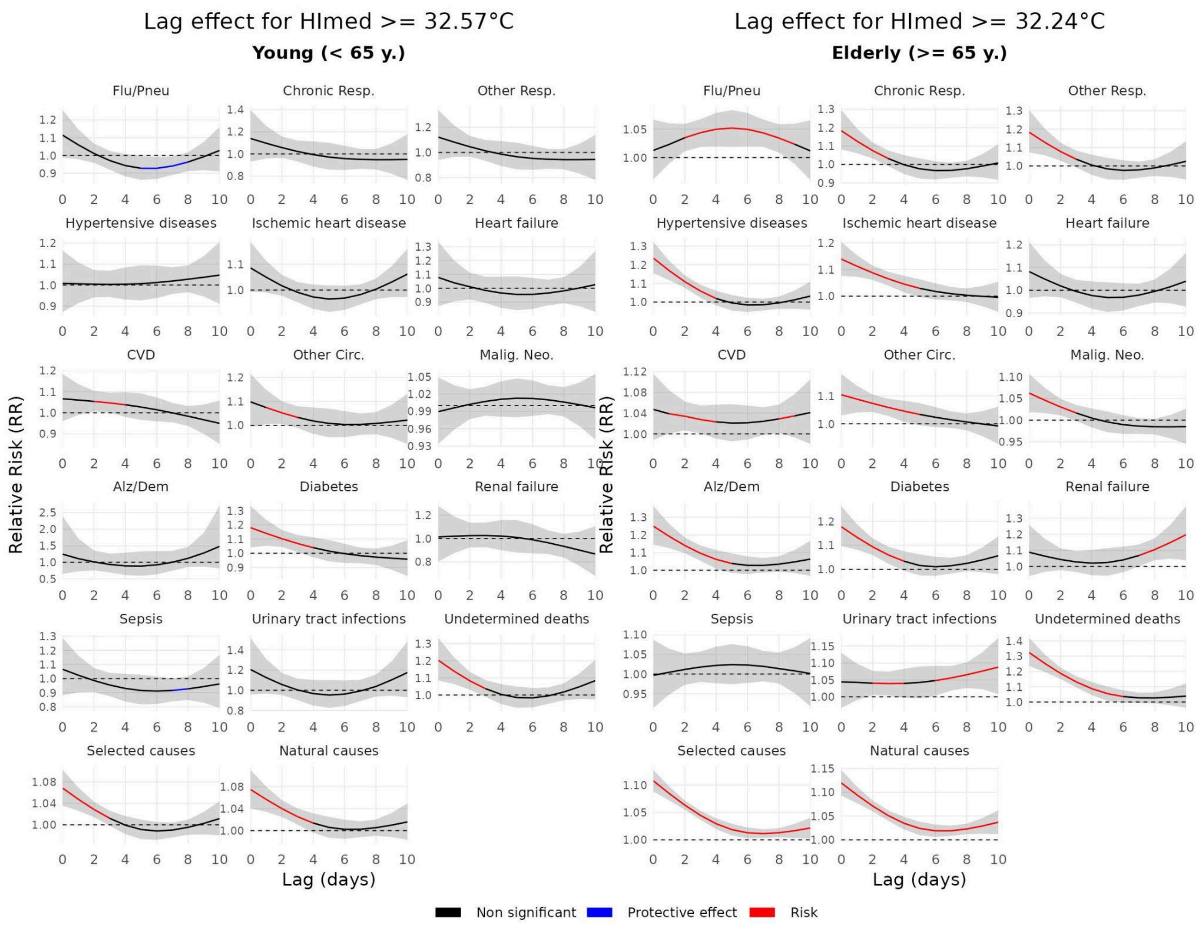


**Figure S6. Mortality Relative Risk (RR) when exposed to high Tmed (RR > 1.25 average threshold) according to lag days, for all causes included among young (left) and elderly (right). RJC, Jan/2012-Jun/2024.**

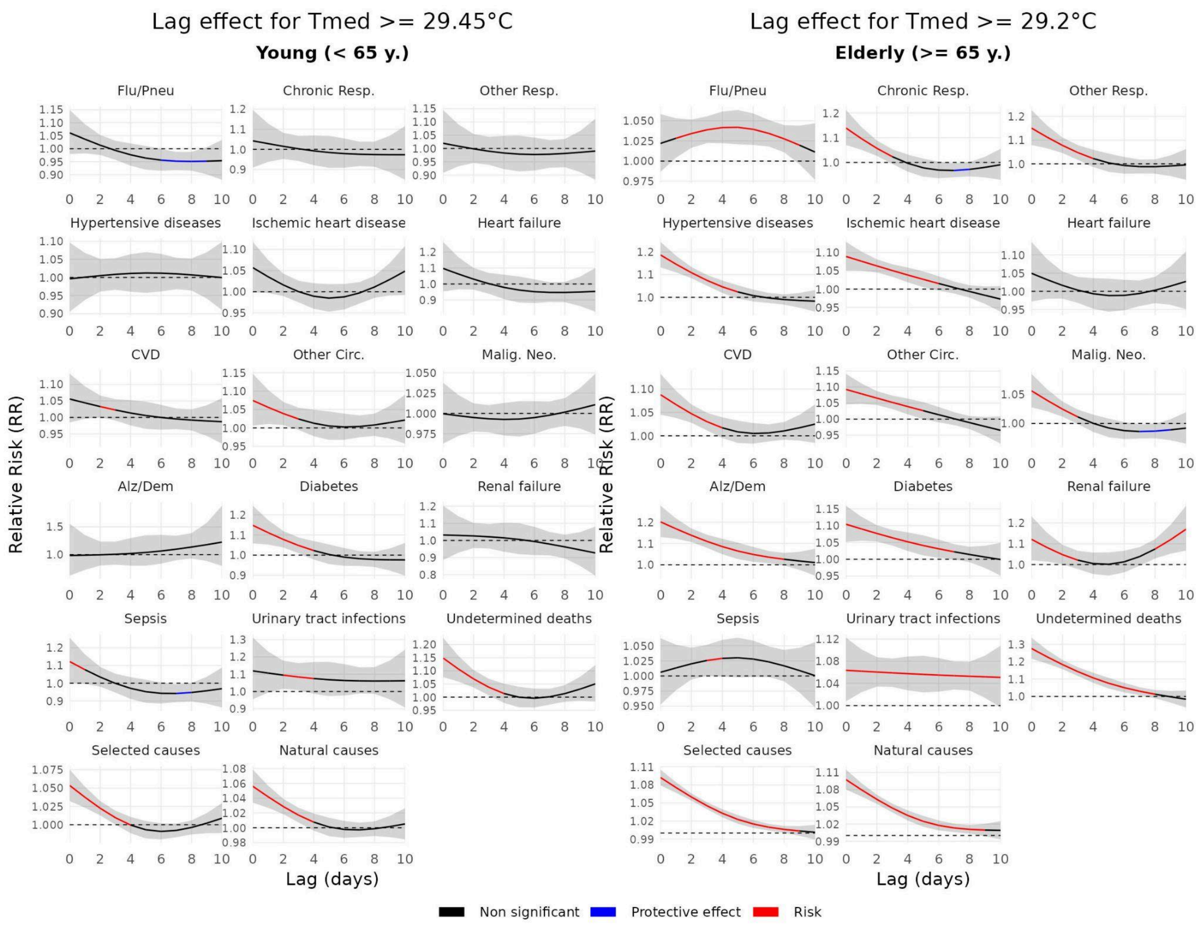


**Figure S7. Best-performing models considering AIC for HAAT variables, varying HI threshold (HI >= 32°C / HI >= 36°C) and accumulated period (1, 3, 5 and 7 days).**

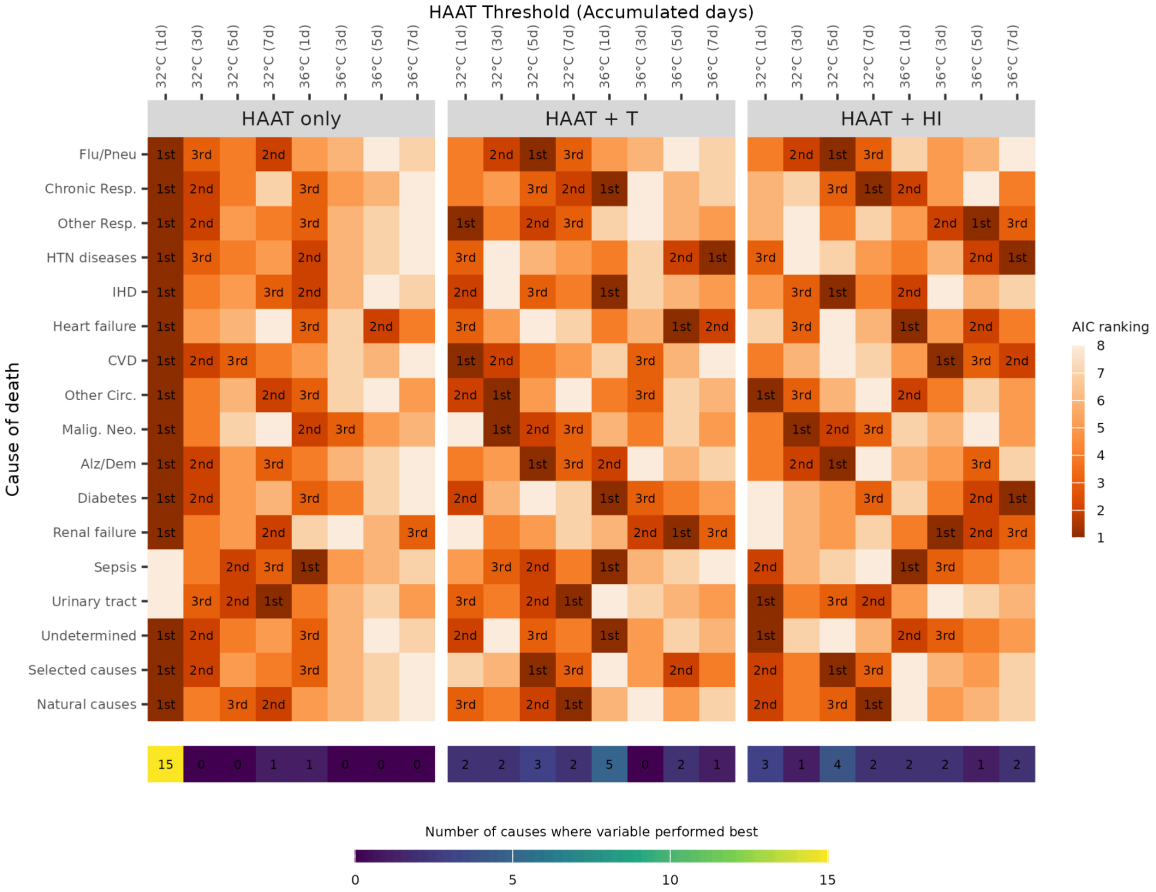


**Figure S8. HAAT values (°C*h) necessary (dots) to reach a relative risk for mortality of 1.25, 1.50 and 2.0, among the elderly. Dashed lines represent the median values through all the causes included.**

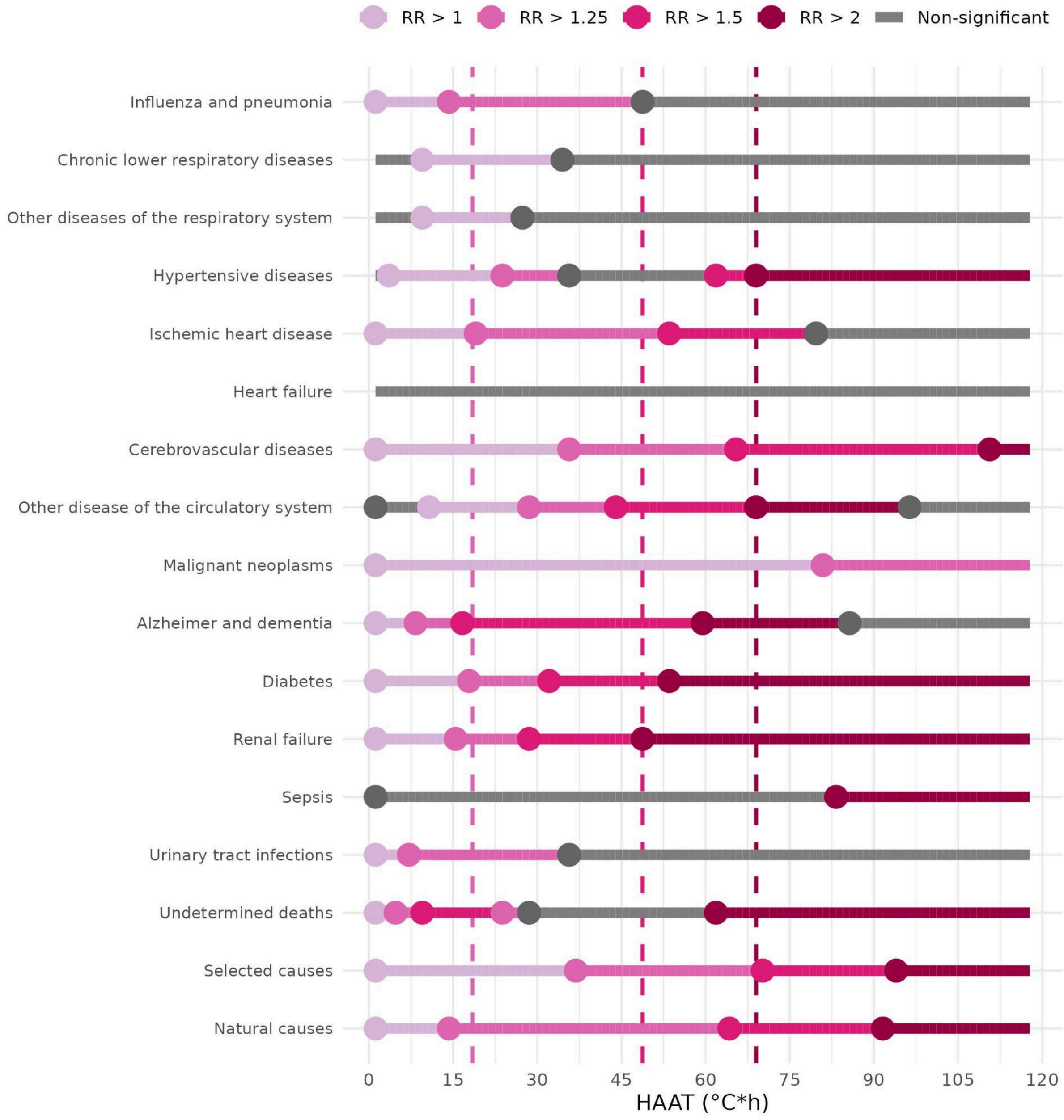


**Figure S9. RJC municipality contour and locations of meteorological stations included in the study, according to data source. Map tiles source: ESRI imagery.**

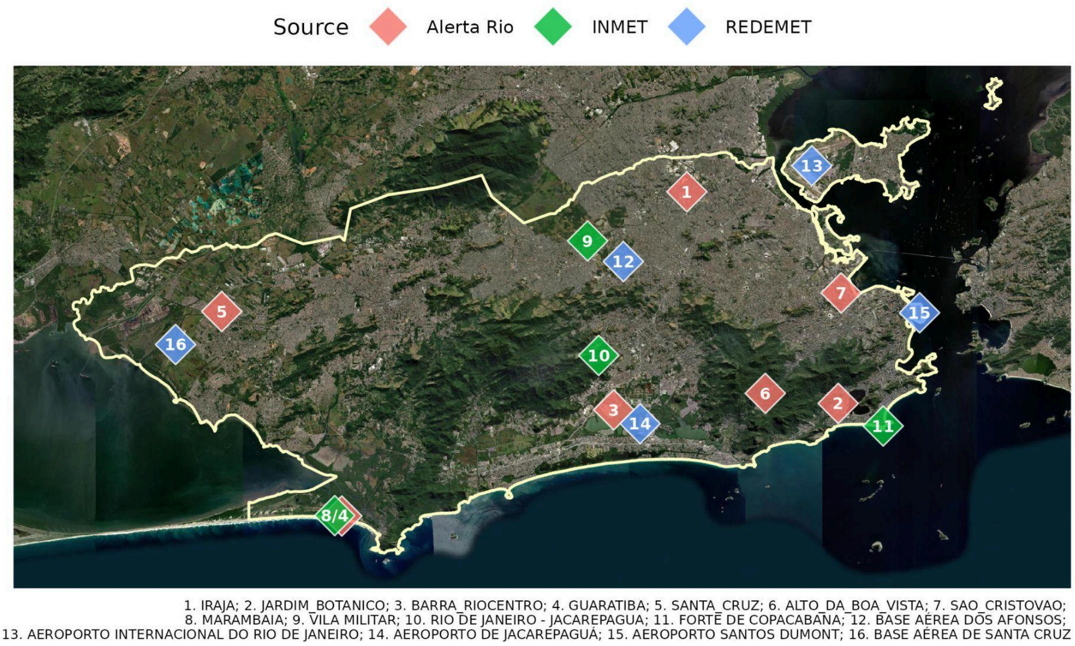

Source
Alerta Rio
INMET
REDEMET
1
2
3
5
6
7
8/4
9
10
11
12
13
14
15
16
1. IRAJA; 2. JARDIM_BOTANICO; 3. BARRA_RIOCENTRO; 4. GUARATIBA; 5. SANTA_CRUZ; 6. ALTO_DA_BOA_VISTA; 7. SAO_CRISTOVAO;
8. MARAMBAIA; 9. VILA MILITAR; 10. RIO DE JANEIRO - JACAREPAGUA; 11. FORTE DE COPACABANA; 12. BASE AÉREA DOS AFONSOS;
13. AEROPORTO INTERNACIONAL DO RIO DE JANEIRO; 14. AEROPORTO DE JACAREPAGUÁ; 15. AEROPORTO SANTOS DUMONT; 16. BASE AÉREA DE SANTA CRUZ
